# Epigenetic Activation of the EBV Protein Kinase Determines Antiviral Drug Response in Central Nervous System Lymphoma

**DOI:** 10.1101/2024.08.17.24311219

**Authors:** Christoph Weigel, Haley L. Klimaszewski, Selamawit Addissie, Sarah Schlotter, Fode Tounkara, James P. Dugan, Bradley M. Haverkos, Lynda Villagomez, Mark Lustberg, Pierluigi Porcu, Timothy Voorhees, Michael A Caligiuri, Ginny Bumgardner, Christopher C. Oakes, Robert A. Baiocchi

## Abstract

Epstein-Barr virus (EBV)-associated central nervous system lymphoproliferative diseases (CNSL) are aggressive clinical conditions with poor prognosis. We have reported that durable responses in patients with primary CNS post-transplant lymphoproliferative disease (PTLD) were associated with detection of two ganciclovir (GCV)/zidovudine (AZT) viral drug target proteins, the EBV kinases BGLF4 and BXLF1 in CNSL biopsies. These are associated with lytic EBV and the mechanism for expression in latently infected EBV+ CNSL has been unknown.

By carrying out RNA expression analysis in CNSL tissue biopsies (n=12), we confirmed expression of *LMP1*, *BXLF1*, and *BGLF4*, but not *BZLF1*, pointing to an incomplete lytic EBV program. Biopsies from systemic PTLD (n=24) were used for comparison and showed significantly less expression of *BGLF4*. By quantifying DNA methylation in EBV gene promoters we showed significantly decreased promoter methylation at *BGLF4* in CNSL versus systemic PTLD (p=0.0006). Luciferase reporter analysis of the *BGLF4* upstream sequence revealed 3 regions of promoter activity and 5ˈ RACE in n=4 EBV-infected cell lines and n=5 CNSL biopsy samples identified transcription start sites at these promoters. We identified DNA methylation loss at single CpG dinucleotides which were specifically demethylated in CNSL, while surrounding EBV methylation remained high. Lastly, TET2 knockout and expression of TET1/2-suppressive mutant IDH1 in a latent HEK293 EBV model indicated that active demethylation is necessary for activity of *BGLF4* promoters. We detail the epigenetic basis of *BGLF4* expression in CNSL via locus-specific promoter activation that may hold value for determination of antiviral drug sensitivity.

**Significance:** We show site-specific DNA methylation loss as the molecular basis of *BGLF4* expression in CNSL. *BGLF4* methylation holds potential for further development as a biomarker of antiviral drug response in EBV+ CNSL.

## Introduction

Epstein-Barr Virus (EBV) is a gammaherpesvirus responsible for a broad spectrum of human diseases including hematologic malignancies (1). EBV maintains lifelong infection by entering a state of latency inside its host cell, silencing many of its genetic elements. EBV can undergo lytic activation, resulting in expression of its numerous lytic genes, viral DNA replication and eventually release of infectious viral particles (2).

Host cellular epigenetic mechanisms of gene expression have been identified as widespread and indispensable for the EBV life cycle (2,3). Among these, DNA methylation primarily serves as a gene silencing mechanism of EBV genetic elements in latency, allowing EBV to suppress its numerous lytic transcripts and evade immune detection (4). Lytic activation necessitates removal of DNA methylation for concerted programs of lytic gene expression (2). The rapid production of EBV DNA copies upon lytic cycle entry leaves most of its genome completely demethylated (2).

EBV lytic/latent molecular mechanisms have emerged as promising avenues to diagnose and treat EBV malignancy (1,5–7). However, current clinical assays for EBV are limited to detecting the presence of EBV via PCR (8,9) and do not provide important information on EBV gene expression or overall lytic/latent activity states. Thus, making clinical decisions based on EBV detection alone has remained challenging.

This is particularly true in states of immunosuppression that predispose patients to both EBV-driven oncogenesis and lytic viral activation, often complicating differential diagnosis. These conditions include iatrogenic immune suppression in the context of organ transplants (post-transplant lymphoproliferative disease, PTLD) (10). Among PTLDs, central nervous system involvement occurs in 10-15% of cases. These CNS lymphoproliferative diseases (CNSL) are overwhelmingly EBV-associated (EBV+) and primary CNSL (PCNSL), which are confined to the CNS (11). CNSL stands out as particularly aggressive, commonly exhibiting infiltrative growth patterns that make resection impractical (12). Patients face significant co-morbidities, limiting current treatment options like methotrexate (13), with drug toxicity as a major concern (14). Additional evidence-based treatment options are needed to improve outcomes for this population (15).

We have previously reported that EBV+ CNSL respond exceptionally well to the GARD treatment regimen comprising Rituximab, dexamethasone and the antivirals ganciclovir (GCV) and zidovudine (AZT) (13). Immunohistochemical staining revealed the presence of viral EBV protein- and thymidine kinases (EBV BGLF4 and BXLF1, respectively) in CNSL biopsies. These EBV proteins are required for specific antiviral drug activation via phosphorylation of GCV and AZT from their prodrug states, thus establishing a mechanistic rationale for their therapeutic efficacy in EBV-infected cells (16,17). The presence of BGLF4 and BXLF1 thus confers unique vulnerabilities to tumor cells that can be exploited for virus-targeted antitumor therapy (7,18). It has remained unclear if there are specific regulatory EBV features that could indicate the presence of BGLF4/BXLF1, and thus serve as potential biomarkers to identify tumors that respond to GCV/AZT treatment.

We set out to identify molecular features of *BGLF4/BXLF1* expression in CNSL. Using data from patient samples and in vitro models, we conclude that epigenetic activation and expression of *BGLF4* is present in otherwise latent EBV, and thus provide a new perspective on locus-specific gene control of EBV lytic genes. We found that *BGLF4* DNA methylation loss is a surrogate of *BGLF4* expression and may be explored as a future biomarker for targeted CNSL treatment with antivirals.

## Materials and Methods

### Clinical Data

Patients with EBV+ CNSL who were treated with the GARD regimen at the Ohio State University (OSU) Wexner Medical Center between January 1998 and June 2024 were selected for this study. Research was approved by the OSU institutional review board record (IRB# 2023C0117) and clinical data was accessed retrospectively from the electronic medical records (Table 1, Table 2). Pathology was reported according to the original diagnosis and pathology reports. Complete response (CR), unconfirmed complete response (CRu), partial response (PR), and progressive disease (PD) were defined using guidelines from the International Primary CNS Lymphoma Collaborative Group (19). The GARD regimen consisted of 14 days of induction treatment with intravenous GCV (5mg/kg twice daily), AZT (1,500mg twice daily), and dexamethasone (10-40 mg). Four doses of rituximab (375 mg/m^2^) were given on days 1, 8, 15, and 22. Following induction, patients were prescribed maintenance oral valganciclovir (450mg twice daily) and AZT (300 mg twice daily).

**Table 1.** CNSL Patient characteristics. n=24 patients with CNSL treated at OSU with GARD are listed. EBV DNA was determined with a clinical diagnostic qPCR assay at OSU Medical Center. DLBCL: diffuse large B-cell lymphoma; LYG: Lymphomatoid Granulomatosis; CSF: cerebrospinal fluid.

| ID | Age Group (years) | Sex | Race | Cause of CNSL | Primary or Secondary | Reason for Immunosuppression | Pathologic Diagnosis | Biopsy EBER Status | EBV DNA in Peripheral Blood | EBV DNA in CSF | Reported previously (13) | DNA methylation cohort |
| --- | --- | --- | --- | --- | --- | --- | --- | --- | --- | --- | --- | --- |
| 1 | 30-39 | M | W | PTLD | Primary | Kidney transplant | Grade 3 LYG | Positive | None detected (<2000 copies/mL) | Detected (no viral load) | Yes | Yes |
| 2 | 40-49 | M | W | PTLD | Primary | Kidney/Pancreas transplant | Grade 3 LYG | Positive | None detected (<2000 copies/mL) | Not tested | Yes | Yes |
| 3 | 20-29 | M | W | PTLD | Primary | Kidney transplant ('97, '04) | DLBCL | Positive | None detected (<2000 copies/mL) | Not tested | Yes | Yes |
| 4 | 40-49 | M | W | PTLD | Primary | Kidney transplant | DLBCL | Positive | Not tested | Not tested | Yes | No |
| 5 | 50-59 | M | W | PTLD | Primary | Kidney transplant | Large B-cell PTLD | Positive | Detected (1500 copies/mL) | None detected | Yes | No |
| 6 | 70-79 | F | W | PTLD | Primary | Kidney transplant | Polymorphic PTLD | Positive | None detected (<2000 copies/mL) | Detected (no viral load) | Yes | No |
| 7 | 50-59 | F | W | PTLD | Primary | Kidney transplant | DLBCL | Positive | Detected (2600 copies/mL) | Detected (555 copies/mL) | Yes | No |
| 8 | 60-69 | F | W | PTLD | Primary | Kidney transplant | DLBCL | Positive | Detected (1000 copies/mL) | Detected (500 copies/mL) | Yes | No |
| 9 | 60-69 | F | B | PTLD | Primary | Liver transplant | B-cell PTLD | EBV+ in CSF | None detected (<2000 copies/mL) | Detected (322,000 copies/mL) | Yes | No |
| 10 | 40-49 | M | W | PTLD | Primary | Kidney/Pancreas transplant | Lymphohistiocytic infiltrate with monoclonal T-cells | Positive | Detected (2286 copies/mL) | Detected (<10,000 copies/mL) | Yes | Yes |
| 11 | 30-39 | M | W | PTLD | Primary | Kidney transplant ('00,'19) | DLBCL | Positive | Detected (7880 copies/mL) | Detected (>10,000 copies/mL) | No | Yes |
| 12 | 70-79 | F | W | Other Immunosuppr. | Primary | Seronegative arthritis | Grade 3 LYG | Positive | None detected (<2000 copies/mL) | Not tested | No | No |
| 13 | 60-69 | M | W | Other Immunosuppr. | Secondary | Epidermolysis bullosa acquisita, HSCT, BMT for NHL | DLBCL | Positive | None detected (<2000 copies/mL) | Detected (>10,000 copies/mL) | No | Yes |
| 14 | 60-69 | M | W | Other Immunosuppr. | Secondary | Psoriasis (MTX) | Large cell lymphoma of ambiguous lineage | Positive | Detected (78,693 copies/mL) | Not tested | No | No |
| 15 | 40-49 | M | B | Sporadic PCNS | Primary | None | B-cell PCNS-LPD | EBV+ in CSF | Detected (1464 copies/mL) | Detected (<10,000 copies/mL) | No | Yes |
| 16 | ≥80 | F | W | Other Immunosuppr. | Primary | Interstitial lung disease | DLBCL | Positive | None detected (<1000 copies/mL) | Not tested | No | No |
| 17 | 70-79 | M | W | PTLD | Secondary | Kidney transplant | DLBCL | Positive | None detected (<2000 copies/mL) | Detected (<10,000 copies/mL) | No | Yes |
| 18 | 70-79 | M | W | PTLD | Primary | Kidney transplant | DLBCL | Positive | None detected (<2000 copies/mL) | Not tested | No | Yes |
| 19 | ≥80 | M | W | Other Immunosuppr. | Primary | Advanced age | Grade 3 LYG | Positive | None detected (<2000 copies/mL) | Not tested | No | Yes |
| 20 | 50-59 | M | Indian | Other Immunosuppr. | Primary | Wegner's granulomatosis and anti-GBM disease | DLBCL | Positive | None detected (<2000 copies/mL) | Not tested | No | Yes |
| 21 | 20-29 | F | Indian | PTLD | Primary | Kidney transplant ('19, '22) | High grade B-cell PTLD | Positive | Unknown (outside hospital) | Detected (47,000 copies/mL) | No | No |
| 22 | 70-79 | M | W | PTLD | Primary | Heart transplant | DLBCL | Positive | Detected (13,695 copies/mL) | Not tested | No | Yes |
| 23 | 70-79 | F | W | Other Immunosuppr. | Primary | Type 2 diabetes | Grade 3 LYG | Positive | None detected (<2000 copies/mL) | Unknown (outside hospital) | No | No |
| 24 | <20 | F | W | Other Immunosuppr. | Primary | Juvenile rheumatoid arthritis | B-cell PCNS-LPD | EBV+ in CSF | None detected (<1000 copies/mL) | Detected (<10,000 copies/mL) | No | No |

**Table 2.** Treatment response and survival in CNSL patients receiving GARD. ARA-C: Cytosine arabinoside or Cytarabine; WBXRT: whole brain radiation; RM-CHOP: Rituximab Methotrexate Cyclophosphamide Doxorubicin Vincristine; RB: Rituximab Bendamustine; Dex: Dexamethasone; PLEX: plasma exchange, IV: intravenous; PO: peroral; BID: twice daily; Survival: A: alive, D: deceased; *patient received GARD for a PTLD lesion in lung.

| ID | Transplant Induction | Immunosuppression at Diagnosis | Prior Treatment | Alterations to Induction | Response to GARD | Additional Treatment | Survival | Response at time of death | Survival time in years |
| --- | --- | --- | --- | --- | --- | --- | --- | --- | --- |
| 1 | ATG | Neoral/cellcept prednisone | None | No Dex | CR | None | A | N/A | 15.4 |
| 2 | ATG | Rapamune/Myfotic | None | No dex, 1 week of 2.5 mg/kg GCV then increased to 5mg/kg | Cru | Repeat induction | D | CRu | 11.5 |
| 3 | ATG/cellcept + pheresis ('04) | Rapamune/Cellcept ('04) | None | None | CR | None | A | N/A | 14.2 |
| 4 | Unk | Cellcept prednisone | None | No rituximab, 2.5 mg/kg GCV | CR | None | D | CR | 11.8 |
| 5 | ATG | Neoral/Myfotic | None | No rituximab, 1.25 mg/kg GCV | CR | None | D | PD | 3.8 |
| 6 | ATG | Neoral/Myfotic | Steroids | 1200mg AZT | CRu | None | D | CRu | 1.7 |
| 7 | Unk | Neoral/cellcept | Rituximab WBXRT | None | CRu | None | A | N/A | 13.3 |
| 8 | ATG | Cellcept/rapamune prednisone | None | Two doses of rituximab | CRu | None | D | PD | 0.3 |
| 9 | Unk | Neoral/Myfotic prednisone | None | None | SD | Rituximab, IV AZT/GCV, WBXRT | D | PD | 0.3 |
| 10 | ATG | Myfotic, tacrolimus | None | 1200mg AZT | CR | None | A | N/A | 10.7 |
| 11 | Unk | Cellcept/rapamune prednisone | None | None | PD | MTX, IV AZT | A | N/A | 10.2 |
| 12 | - | Methotrexate | None | None | CR | None | A | N/A | 10.0 |
| 13 | Etoposide, ARA-C, Platinum | Cellcept prednisone | None | IV AZT/GCV for 13 days, rituximab delayed | PR | PO AZT/GCV | D | PD | 0.9 |
| 14 | - | Adalimumab | RM-CHOP, Brentuximab, Romidepsin, BR | All treatments stopped after 1 week | PD | None | D | PD | 0.05 |
| 15 | - | - | None | None | CR | None | A | N/A | 3.3 |
| 16 | - | Cellcept | Dex., PLEX | None | SD | None | D | SD | 0.2 |
| 17 | ATG | Everolimus Neoral | GARD* | None | PR | MTX, Rituximab, Dex | D | CR | 5.4 |
| 18 | ATG | Rapamune Myfotic | None | AZT stopped at one week | CR | None | D | CR | 2.2 |
| 19 | - | - | None | None | CRu | None | A | N/A | 6.4 |
| 20 | - | Cellcept<br>prednisone | None | IV medications held on days 9,<br>11, 12, 13 due to mental status<br>change and refusal | PR | Rituximab and<br>MTX | D | CRu | 1.2 |
| 21 | Unk | Cellcept | None | AZT PO | CRu | None | A | N/A | 2.6 |
| 22 |  | Neoral/cellcept<br>prednisone | None | 2.5 mg GCV,<br>1200mg AZT | PD | None | D | PD | 0.1 |
| 23 | - | - | None | None | PR | None | D | PD | 0.8 |
| 24 | - | Infliximab | None | None | CR | None | A | N/A | 5.1 |

### Sample Collection and Consent

Tissue samples at diagnosis were obtained from the OSU Tissue Archive Services at the Department of Pathology. Patients provided consent to research via Ohio State University institutional review boards (IRB) protocols. Donor lymphoblastoid cell lines (LCLs) were generated from peripheral blood mononuclear cells (PBMCs) from EBV-seropositive healthy individuals with donor consent as described previously (20).

### DNA and RNA isolation

Tissue biopsy DNA and RNA was isolated with QIAamp DNA FFPE Tissue Kit (QIAGEN) and RNeasy FFPE Kit (QIAGEN) according to manufacturer instructions. DNA and RNA from cell culture was isolated with QIAamp DNA Mini Kit (QIAGEN) and TRIzol reagent (Invitrogen), respectively. Plasmid and lentiviral vector DNA were prepared with ZymoPURE Plasmid Miniprep Kit (Zymo Research) and PureYield Plasmid Midiprep System (Promega).

### Cell culture

Cells were grown in a humidified cell culture incubator at 5% CO2 and 37°C. All LCLs and hematopoetic cell lines were maintained in RPMI 1640 with 10% (v/v) fetal bovine serum (FBS) supplemented with penicillin/streptomycin (Gibco) and GlutaMAX (Gibco). LCLs were generated as described (20). HEK293, MEC1/2 and Raji lines were obtained from Christoph Plass’s group (German Cancer Research Center, Heidelberg, Germany). YT cells were obtained from Bethany Mundy’s group (The Ohio State University, Columbus, OH). Lenti-X HEK293T cells were obtained from Takara Bio. M81 EBV-carrying HEK293 cells (21) were a gift from Henri-Jaques Delecluse’s group (German Cancer Research Center, Heidelberg, Germany). HEK293/293T cells were maintained in DMEM media (Gibco) with 10% (v/v) FBS, penicillin/streptomycin and GlutaMAX. Burkitt lymphoma lines Kem, Mutu and Rael were obtained from Lisa Giulino-Roth’s group (Weill Cornell Medical College, New York, NY). All cell lines were confirmed mycoplasma-free via the MycoAlert Mycoplasma Detection Kit (Lonza) every two weeks during experiments.

### Lentiviral transduction

IDH1 R132H plasmid (gift from Eric Campeau (22), Addgene plasmid #81686) was cloned into pLenti-PGK Blasti DEST lentiviral vector (gift from Jesse Boehm (23), Addgene plasmid #19065) using the Gateway cloning system (Invitrogen) in One Shot Stbl3 Chemically Competent E. coli (Invitrogen). Lentivirus particles were produced in HEK293T cells using standard methods. Viral supernatant was concentrated using Lenti-X Concentrator solution (Takara Bio) following the manufacturer protocol. Concentrated lentivirus preparations were mixed with 1.0e6 live HEK293 cells to yield a MOI of 5. Cells were selected with Blasticidin S HCl (10 µg/ml, GoldBio) for at least 14 days.

### CRISPR-Cas9 gene knockout model

*TET2* knockout was achieved with CRISPR-Cas9-mediated gene disruption using the TrueTag system (Invitrogen). Two homology templates (Puromycin/Blasticidin resistance-CMV-GFP/RFP) were designed together with a guideRNA targeting the *TET2* locus in both alleles with the TrueDesign Genome Editor software (Invitrogen) and PCR-amplified. Homology templates (1250ng each) were mixed with 6.2µg of TrueCut HiFi Cas9 Protein (Invitrogen), 1200ng of guideRNA (Invitrogen, Suppl. Table 2) and 5e6 HEK293 cells in 100µl total volume of Electroporation buffer (MaxCyte) and electroporated using a MaxCyte GTx device (MaxCyte). Cells were selected under Puromycin Dihydrochloride (1µg/ml, Gibco) and Blasticidin S HCl (10µg/ml, GoldBio) for at least 14 days. GFP/RFP double-positive cells were then sorted on a FACSAria device (BD Biosciences) and maintained under Puromycin and Blasticidin.

### qPCR and qRT-PCR

EBV was detected in DNA samples via primers amplifying the *EBNA1* and normalized to human genome equivalents (*ACTB* locus, Suppl. Table 2), using PCR with Fast SYBR Green Master Mix (Applied Biosystems). RNA expression of EBV genes was assessed using quantitative reverse-transcriptase PCR (qRT-PCR). 1µg of total isolated RNA was reverse-transcribed using Superscript IV reverse transcriptase (Invitrogen) according to manufacturer protocol with random hexamer oligonucleotides (QIAGEN). qPCR with primers for EBV *BZLF1*, *BXLF1*, *BGLF4* and *LMP1* was carried out and normalized to human *ACTB* signal (Suppl. Table 2). qRT-PCR was performed using TaqMan Fast Advanced Master Mix (Applied Biosystems) and PrimeTime qPCR Probe Assays with FAM/ZEN/IBFQ probes (Integrated DNA technologies). Samples were run in technical duplicates and only included in analysis when both readouts yielded gene expression values within 15% range of each other. qPCR was performed on a QuantStudio 7 Pro real-time qPCR system (Applied Biosystems).

### Western Blot

Western Immunoblots were carried out with standard methods using SDS-PAGE (Bio-Rad). Samples were blotted on PVDF membranes (Bio-Rad) and antibodies against IDH1 (Thermo Scientific, Cat.-N.: A304-161A-T), IDH1 R132H (Thermo Scientific, Cat.-No.: Z2010RT), TET2 (Thermo Scientific, Cat.-No.: A304-247A-T), GFP (ABclonal, Cat.-No.: AE012) and ACTB (Cell Signaling Technologies, Cat.-No.: 8H10D10) as loading control. Signals were visualized on an Odyssey CLx (LI-COR Biotech) using IRDye 800CW goat-anti-rabbit and IRDye 680RD goat-anti-mouse secondary antibodies (LI-COR Biotech).

### DNA methylation assays

DNA methylation analysis of the *LMP1, BGLF4, BXLF1* and *BZLF1* regions (Suppl. Table 2) was carried out using the EpiTYPER assay as described (5). In short, DNA was bisulfite converted using EZ DNA methylation kit (Zymo Research) and EBV regions were PCR-amplified with primers specific for bisulfite DNA (Suppl. Table 2). PCR products were in-vitro transcribed and fragmented with RNase A. Fragments were analyzed via Matrix-Assisted Laser Desorption/Ionization-Time of Flight (MALDI-ToF) mass spectrometry (Agena Bioscience). Ratios of unmethylated versus methylated mass peaks were used to calculate the percentage of DNA methylation.

Global EBV DNA methylation analysis was carried out using the methylation-iPLEX assay as described (24). iPLEX capture and extension primers were designed to amplify and target EBV CpGs from bisulfite-converted DNA sequences using the Typer4.0 software (Agena Bioscience, Suppl. Table 2). All Samples were dispensed onto 384-well SpectroCHIP arrays using the RS1000 Nanodispenser and analyzed using the MassARRAY Analyzer4 system (Agena Bioscience).

### EBV infection

For production of EBV viral particles, cells were co-transfected with EBV *BZLF1*, *BRLF1* and *BALF4* (gp110) mammalian expression plasmids described before (21). Virus collection and EBV infection was carried out with lentiviral transduction protocols described in the methods. After infection, cells were selected with Hygromycin B (200µg/ml, GoldBio) for at least 14 days.

### Luciferase reporter assays

Luciferase assays were carried out in reporter vector pCpG-free promoter-lucia (Invivogen) as published previously (25). *BGLF4* fragments were PCR amplified (Suppl. Table 2) and cloned into the HindIII/SdaI restriction sites using standard techniques. Reporter vectors were *in-vitro* methylated using M.SssI CpG methyltransferase (Thermo Fisher Scientific). Luciferase reporters were transfected into HEK293 cells with TransIT-LT1 reagent (Mirus Bio), and luciferase activity was read out after 48h in a Synergy HT plate reader (Biotek).

### 5’RACE

*BGLF4* 5’RACE was carried out based on published protocols (26) with modifications. 5µg of total RNA was reverse-transcribed using GSP-RT primer (Suppl. Table 2), poly-A tailed and amplified using nested PCRs with GSP (gene specific), Qt, Qo (outer) and Qi (inner)-primers (Suppl. Table 2 and (26)) with Q5 DNA polymerase (New England Biolabs). Products were gel-purified and cloned into pDONR221 (Invitrogen) with Gateway BP clonase II (Invitrogen) followed by Sanger sequencing at the OSU Genomics Shared Resource. EBV sequences were mapped to the EBV reference genome (RefSeq ID NC_007605.1) to determine 5’ends as transcription start sites.

### Data Analysis and Statistics

Statistical significance was determined with unpaired, two-tailed t-test unless stated otherwise. Receiver-operating characteristics (ROC) curves and sensitivity/specificity were determined with easyROC ver. 1.3.1. Unsupervised clustering was carried out with Cluster3 (27) Using Euclidean Distance and visualized with Treeview. EBV genomic data tracks were generated from EBV reference genome (Refseq ID: NC_007605.1) using Integrative Genomics Viewer. Violin Plots were generated with BoxPlotR (28), with center lines showing medians, box limits indicating the 25th and 75th percentiles. Overall survival (OS) was defined as the time from start of treatment until death from all causes while progression free survival (PFS) was defined as the time from start of treatment until disease progression. OS and PFS were visualized with GraphPad Prism 10. Treatment related mortality (TRM) and was defined as death occurring within ninety days of completion of GARD regimen that is not due to progressive disease.

## Results

### Long-term follow up reveals durable responses to GARD treatment of EBV+ CNSL

A total of n=24 patients with EBV+ CNSL were treated at OSU with GARD between 1998 and 2024, with a subset previously described (13). Here we report additional epigenetic studies and follow-up on these patients. This cohort included patients with PTLD PCNSL as well as patients on iatrogenic immune suppression for autoimmune conditions, secondary PTLD CNSL with peripheral PTLD involvement and two EBV+ PCNSL cases with no evidence of immunosuppression (Table 1). The mean age at diagnosis was 56±19 years (Suppl. Table 1). EBV tumor status was verified via *EBER1* staining of biopsy tissue or detectable EBV viral load in the cerebrospinal fluid in cases where a biopsy was unavailable. Cytopenia and other side effects like nausea and weight loss generally improved by holding induction or reducing maintenance therapy. Diffuse large B-cell lymphoma was the most common histology (n=11; 45.8%) followed by grade 3 lymphomatoid granulomatosis (LYG) (n=5; 20.8%) and other B-cell lymphoproliferative disease (n=5;20.8%). Median survival time was 5.4 years, and n=15 patients (63%) achieved a complete response (CR) or CRu, with an overall response rate of 79%. Two-year survival was 63% (95% CI 46-85%) and five-year survival was 53% (95% CI 37-78%) (Figure 1, Table 2). We show rapid resolution of CNSL lesions upon GARD induction (Fig. 1A). Long-term follow up over 15 years post-treatment revealed durable responses and favorable overall and progression-free survival in CNSL patients treated with GARD (Fig. 1B,C).

**Figure 1.**
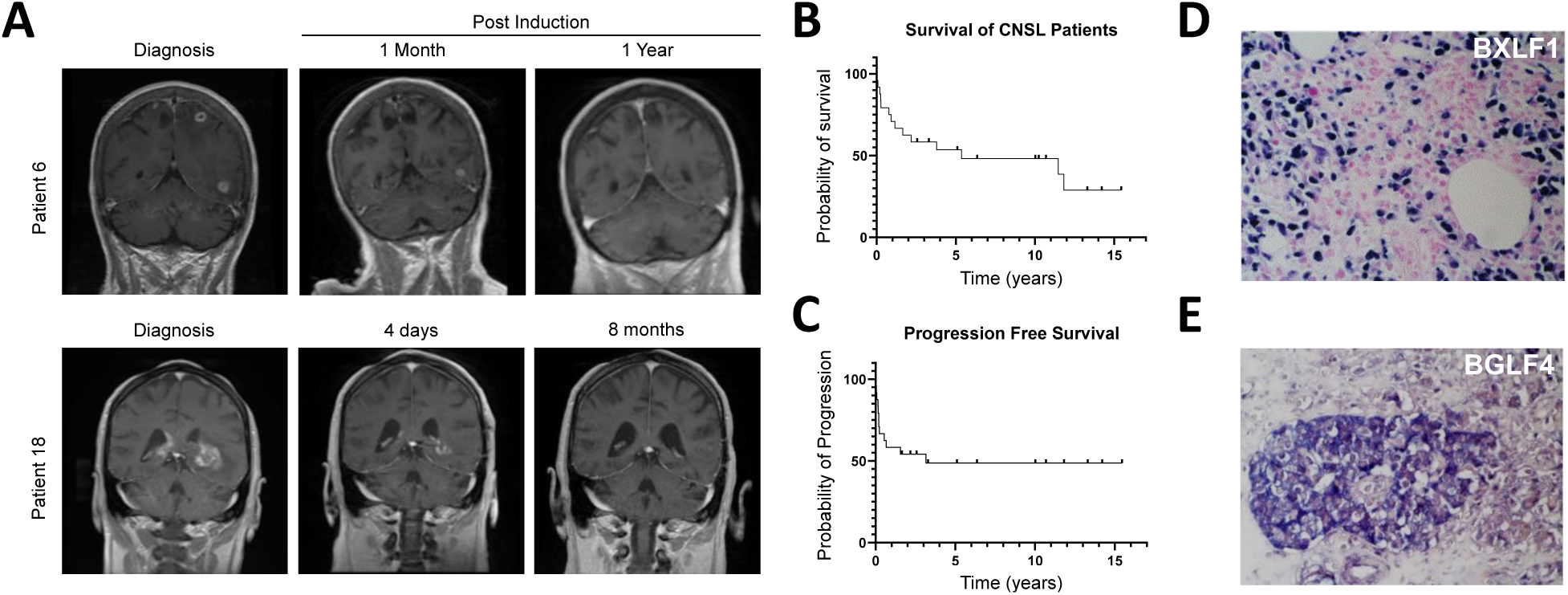
Clinical GARD response characteristics of EBV+CNSL. (**A**) Contrast MRIs from two patients with sequential imaging. Both patients achieved complete and durable that persisted until the patients passed from causes unrelated to PTLD or treatment. (**B,C**) Kaplan-Meier curves representing overall survival (B) and progression-free survival (B) in n=24 CNSL patients treated at OSU. (**D,E**) Representative IHC images for lytic EBV proteins BXLF1 (D) and BGLF4 (E).

### DNA methylation loss in *BGLF4* is associated with *BGLF4* expression in CNSL

We have reported widespread expression of the GCV/AZT target kinases in CNSL tumor biopsies (Fig. 1D,E and (13)). However, the molecular determinant for viral kinase expression remained unknown. In particular, it has remained uncertain if EBV BXLF1/BGLF4 presence was due to lytic replication of EBV. In order to address this question, we carried out gene expression analysis of lytic and latent EBV transcripts in available tumor tissue biopsies of CNSL (n=12, Table 1) and peripheral PTLD patients (n=24, Suppl. Table 1). *BGLF4* showed expression in CNSL significantly above that of peripheral PTLDs, with *BXLF1* expression differences close to statistical significance (Fig. 2A, Suppl. Fig 2A). RNA expression of the essential immediate-early lytic transcript *BZLF1* (Suppl. Fig. 2B), an indicator of lytic cycle entry (2) showed that this transcript was undetectable in many peripheral PTLDs (9/21, 41%) and all but one CNSL (11/12, 92%). The remaining biopsies displayed markedly low *BZLF1*, with the exception of a subset of peripheral PTLDs (6/21, 27%). RNA expression of the EBV latent membrane protein 1 (*LMP1*, Suppl. Fig. 2C) was detectable in the majority of peripheral PTLDs (16/21, 73%) and CNSLs (9/12, 75%), indicating latent EBV.

**Figure 2.**
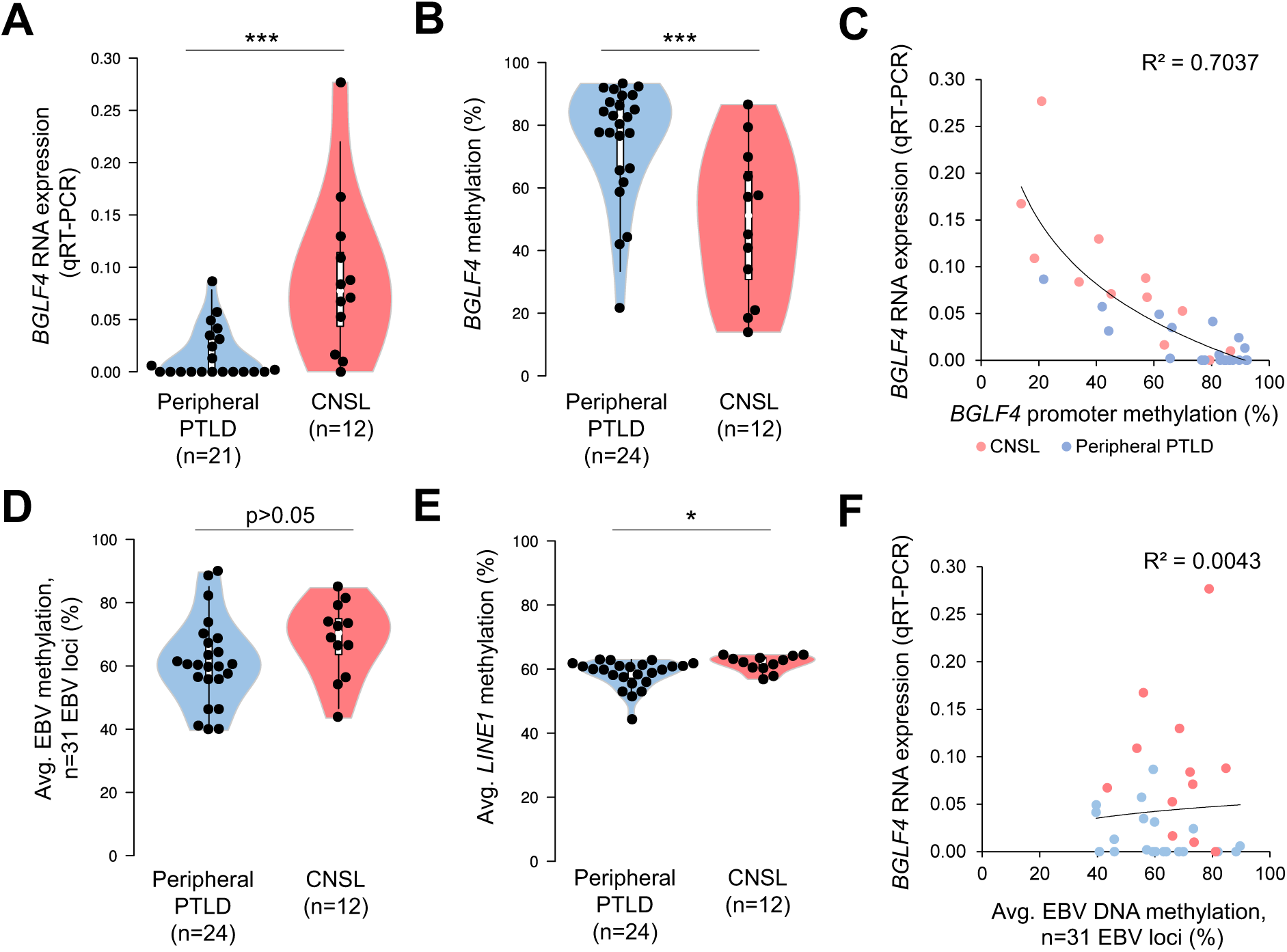
Lowered EBV *BGLF4* DNA methylation and increased *BGLF4* expression in CNSL. (**A**) RNA expression (qRT-PCR) and (**B**) EBV *BGLF4* DNA methylation determined via EpiTYPER in peripheral PTLD and CNSL tissue biopsies. (**C**) correlation of *BGLF4* RNA expression and DNA methylation. (**D**) Average EBV DNA methylation determined via iPLEX assay. Data points show average methylation across 31 EBV CpG sites. (**E**) Average *LINE1* DNA methylation determined via EpiTYPER. (**F**) correlation of *BGLF4* RNA expression and average EBV DNA methylation from (D), R^2^: correlation coefficient. *p<0.05, ***p<0.001, t-test (B,D,E: Exact Wilcoxon-Mann-Whitney test)

To distinguish the specific epigenetic state of EBV loci, we next carried out quantitative DNA methylation profiling of *BGLF4*, *BXLF1*, *BZLF1* and *LMP1* upstream sequences using the EpiTYPER assay (29) in tissue biopsy DNA. We revealed significantly lowered *BGLF4* methylation in CNSL compared to peripheral PTLD (p= 0.0006; Fig. 2B, Suppl. Fig. 2D), but no significant differences in BXLF1, *BZLF1* or *LMP1* (Suppl. Fig. 2D-F), highlighting that these EBV loci have distinct DNA methylation states. Comparison of RNA expression and methylation showed inverse correlation for *BGLF4* (Fig. 2C). We next sought to determine whether the significant EBV methylation differences between CNSL and peripheral PTLDs were the result of global methylation alterations. For this purpose we read out n=31 distinct methylation sites evenly distributed in the EBV genome to establish a representative global EBV methylation state (Fig 2D, Suppl. Fig. 2G). We also read out human genomic *LINE1*, *LTR* and *Alu* repeat element methylation as a surrogate of global host cellular methylation (Fig. 2E, Suppl. Fig. 2H,I). With the exception of a low (avg. 3.2%) methylation difference for *LINE1*, results indicate that global EBV and host cellular DNA methylation are not significantly different in CNSL versus peripheral PTLD, thus indicating a site-specific regulation at *BGLF4* and *BXLF1*. In line with this observation, we found no significant correlation between *BGLF4* mRNA expression and average EBV methylation (Fig. 2F). Lytic EBV activation is characterized by a rapid expansion of EBV DNA copies within a cell, loss of global EBV DNA methylation (2) and widespread lytic gene expression including *BGLF4* (30). We assessed these lytic EBV characteristics by correlating *BGLF4* expression with EBV DNA loads determined in tissue samples via qPCR (Suppl. Fig. 2J) and clinically determined EBV qPCR from serum in peripheral PTLDs (Suppl. Fig. 2K). These comparisons showed no evident correlations, indicating again that *BGLF4* expression is not associated with overall EBV lytic activation. We also assessed the correlation of overall EBV methylation and EBV load (Suppl. Fig. 2L) and again saw no correlation between these readouts. Together, these observations revealed strikingly low DNA methylation at *BGLF4* in CNSL. This lowered methylation did not align with correlates of overall methylation or lytic virus activity (EBV load, EBV methylation, *BZLF1* expression), thus indicating a highly site-specific methylation event while retaining multiple features of viral latency.

### Low DNA methylation at three individual *BGLF4* upstream loci

In order to understand the specificity of lowered methylation in CNSL, we carried out a detailed assessment of DNA methylation in an approximately 1 kilobase upstream region of *BGLF4* previously shown hold transcription initiation sites (TSS) (31). Using the EpiTYPER assay, we derived quantitative, single CpG resolution methylation maps for our CNSL and peripheral PTLD biopsies (Fig. 3A, Suppl. Fig. 3A). We found that significant methylation reduction in CNSL is localized specifically to three differentially methylated regions (DMRs), with each DMR having at least one single CpG dinucleotide with pronounced methylation loss when comparing CNSL and peripheral PTLD (Fig. 3B). Localized *BGLF4* methylation loss was also observed in a subset of peripheral PTLDs (Suppl. Fig 3A), with over one third (9/24, 38%) having at least one DMR with a methylation level below 50%. Comparison of histologic subtypes in CNSL and peripheral PTLDs highlighted a predominance of Diffuse Large B cell lymphoma (DLBCL)-type PTLDs, with significant methylation differences at *BGLF4* CpGs retained when comparing CNS- and peripheral DLBCLs only (Suppl. Fig. 3B-D). All three DMR CpGs displayed inverse correlation with *BGLF4* RNA expression (Fig. 3C-E), thus serving as correlates of the initially observed differential methylation. When categorizing PTLD tissue samples based on the number of hypomethylated (<50% methylation) *BGLF4* DMRs, we found that *BGLF4* RNA expression increased with the number hypomethylated CpG sites (Fig. 3F). Presence of two or more *BGLF4* DMR CpGs below 50% methylation called high (4^th^ quartile) *BGLF4* expression with high sensitivity and specificity (Suppl. Fig. 3E). In summary, methylation analysis of the *BGLF4* upstream region pointed to highly site-specific methylation reduction associated with *BGLF4* expression.

**Figure 3.**
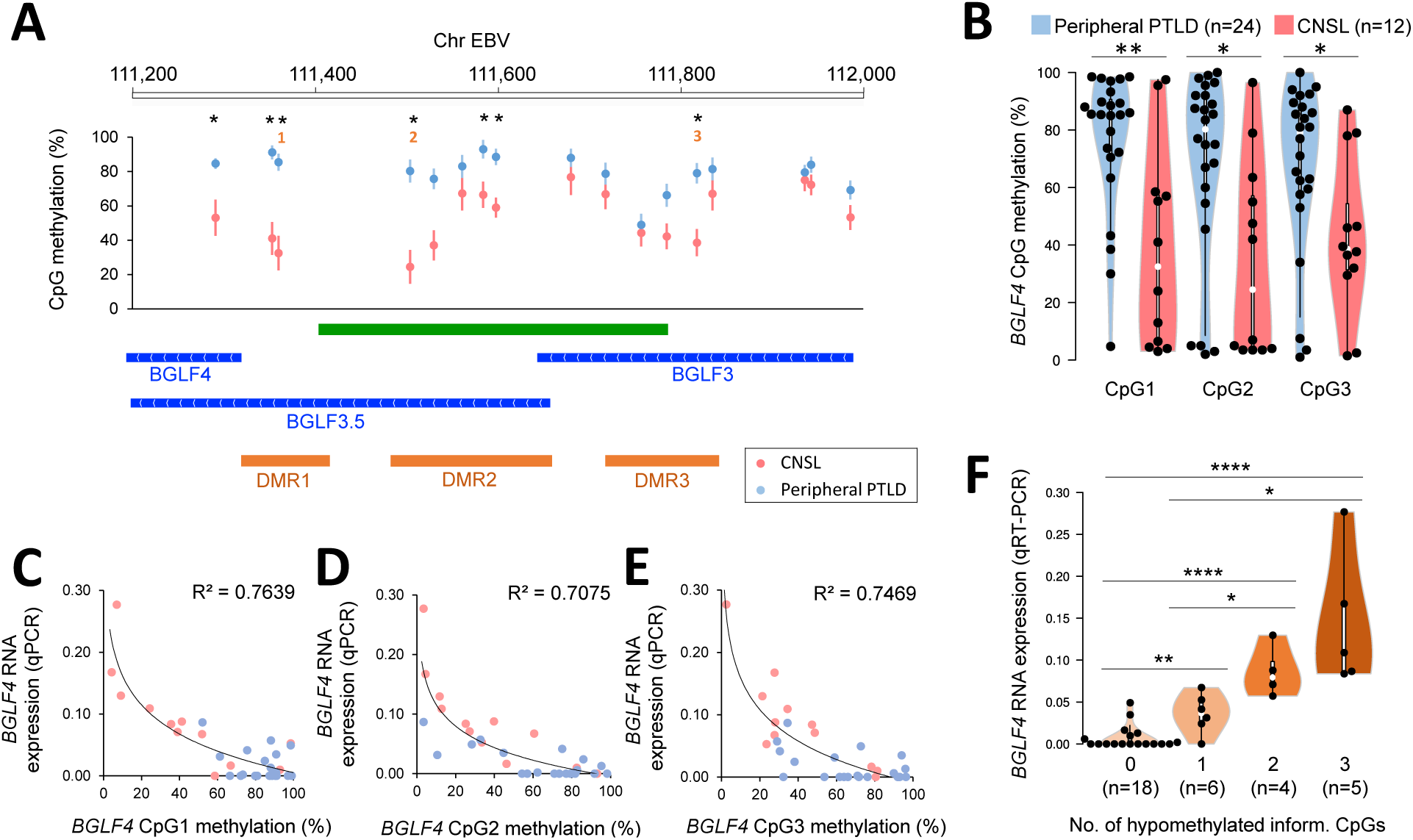
Site-specific methylation loss at EBV *BGLF4*. (**A**) Expanded methylation analysis of the *BGLF4* upstream locus using EpiTYPER assay. EBV genomic coordinates and statistical significance for individual CpGs are annotated above. Data show mean±s.e.m. DNA methylation in CNSL (n=12) and peripheral PTLDs (n=24) cohorts. Lower panel: CpG islands (green), EBV transcripts (blue) and DMRs (orange) are annotated. (**B**) DNA methylation at three individual DMR CpGs marked in (A). (**C-E**) correlation of DNA methylation and *BGLF4* RNA expression in three DMR CpGs of the *BGLF4* upstream locus. (**F**) Classification of *BGLF4* methylation based on number of DMRs with lowered methylation and corresponding *BGLF4* RNA expression. *p<0.05, **p<0.01, ****p<0.0001, t-test. (A,B: Exact Wilcoxon-Mann-Whitney test).

### *BGLF4* DMRs have gene promoter activity and mark three distinct *BGLF4* transcription start sites

The identification of three DMRs upstream of the *BGLF4* open reading frame and their association with gene expression pointed to the presence of gene regulatory elements in that region. We tested the functional properties of *BGLF4* upstream elements in a dual luciferase reporter assay and found that all three DMR regions carried gene promoter activity significantly above that of a control minimal promoter (Fig. 4A). These activities were frequently increased when utilizing a HEK293 model carrying latent EBV (Suppl. Fig. 4A). We confirmed the influence of DNA methylation on promoter activity by using *in vitro* CpG-methylated reporter constructs. DNA methylation significantly decreased DMR promoter activity (Fig. 4B). In order to identify whether the observed regions of promoter activity give rise to RNA transcripts, we next carried out 5’ rapid amplification of cDNA ends (RACE). Using amplification of cDNA ranging from the *BGLF4* start codon into upstream sequences, we identified three distinct *BGLF4* TSS within DMR2 and DMR3 in n=5 primary CNSL samples (Fig. 4C-G) and n=4 EBV cell models (Suppl. Fig. 4B-E, Suppl. Table 3). Combining data from our EBV cell models and CNSL biopsies, we showed that TSS activity was strongly associated with CpG methylation immediately adjacent each TSS (Fig. 4H). This association was confirmed in all three TSS sites individually (Suppl. Fig. 4F) and when including methylation in a 25bp window around TSS (Suppl. Fig. 4G). Together, our data revealed promoter activity in *BGLF4* upstream DMR elements that give rise to distinct viral *BGLF4* transcripts in CNSL.

**Figure 4.**
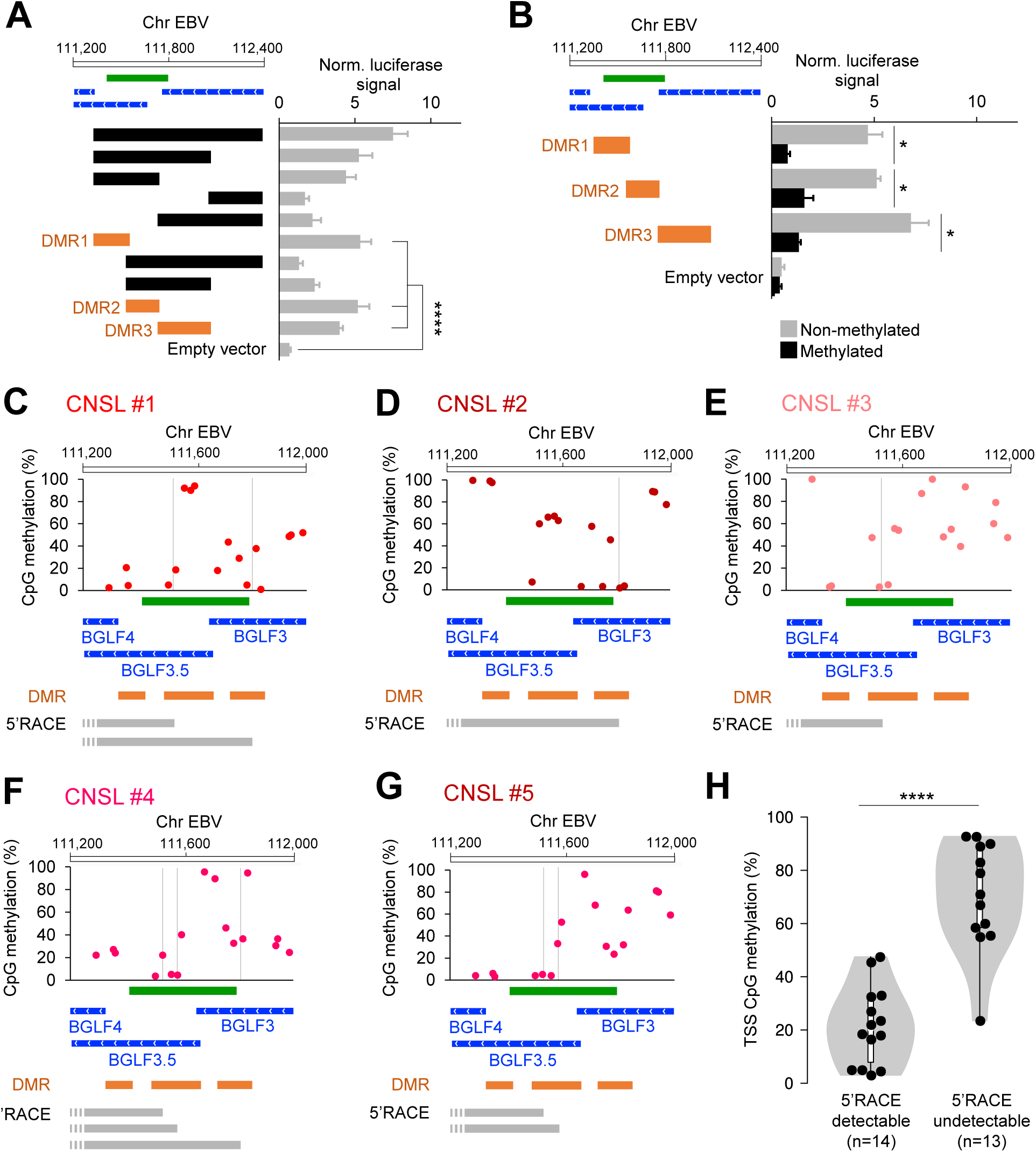
*BGLF4* DMRs possess gene promoter activity. (**A,B**) Luciferase reporter assay of *BGLF4* upstream DNA elements alone (A) or after *in vitro* DNA methylation or reporter constructs (B). DNA sequences representing DMRs are marked in orange. EBV genomic coordinates, CpG islands (green) and EBV transcripts (blue) are indicated above. Bar graphs show mean±standard deviation of quadruplicate readouts in HEK293 cells. (**C-G**) Results from 5’RACE in n=5 CNSL samples with high *BGLF4* RNA expression. Plots show DNA methylation across the *BGLF4* upstream region in each sample with detected TSS marked by vertical lines. Lower panels: CpG islands (green), EBV transcripts (blue), DMRs (orange) and sequenced transcripts from 5’RACE (grey). (**H**) Violin plot depicting DNA methylation at CpG sites closest to TSS with either detectable (n=14) or undetectable (n=13) transcript. Data are aggregate readouts from n=5 CNSL samples from (C-G) and an additional n=4 EBV+ cell lines (Suppl. Fig. 4B-E). *p<0.05, ****p<0.0001, t-test (H: Exact Wilcoxon-Mann-Whitney test).

### TET activity is required for the establishment of lowered *BGLF4* DMR methylation and *BGLF4* expression

We next sought to determine which host epigenetic factors may be required for the establishment of *BGLF4* DNA methylation patterns. Site-specific loss of DNA methylation in the human genome is mediated by the ten-eleven translocation (TET) family of DNA demethylases (32). TET2 was reported to have profound impact on EBV latency and global gene expression patterns (33,34). To test whether TET2 may also be required for methylation regulation at *BGLF4*, we implemented a model that recapitulates the establishment of DNA methylation patterns on EBV following host cell infection and latency establishment. For that we used HEK293 cells and the epitheliotropic EBV strain M81 (Suppl. Fig. 5A). We tested the impact of biallelic CRISPR knockout of *TET2* in that setting (Suppl. Fig. 5B). We again used the EpiTYPER assay to interrogate DNA methylation at *BGLF4*. Comparison of EBV infection in control or biallelic *TET2* knockout (TET2−/−, Fig. 5A) revealed that a region around the active *BGLF4* TSS significantly gained DNA methylation in a TET2−/− setting (Fig. 5B). This coincided with significantly decreased *BGLF4* RNA expression (Fig. 5C). We found that TET2 knockout did not significantly alter the global EBV methylation average (Fig. 5D, Suppl. Fig. 5C). We next validated these findings by establishing a second model of TET deficiency that relies on overexpression of mutant isocitrate dehydrogenase 1 (IDH1 R132H) which impairs all three TET family members by generating an alternative metabolite, alpha-hydroxyglutatrate (35). Based on this mechanism, IDH1 R132H provides a distinct mode of TET inhibition that targets enzymatic activity directly. We found that lentiviral overexpression of IDH1 R132H (Suppl. Fig. 5D) followed by EBV M81 infection in HEK293 cells elevated DNA methylation and lowered RNA expression at *BGLF4* (Suppl. Fig. 5E-G). In contrast to *TET2* knockout, IDH1 R132H expression also resulted in a modest but significant increase in global EBV methylation (Suppl. Fig. 5H,I). Taken together, we deployed two distinct models of TET deficiency and found that the site-specific decrease of *BGLF4* methylation requires TET2.

**Figure 5.**
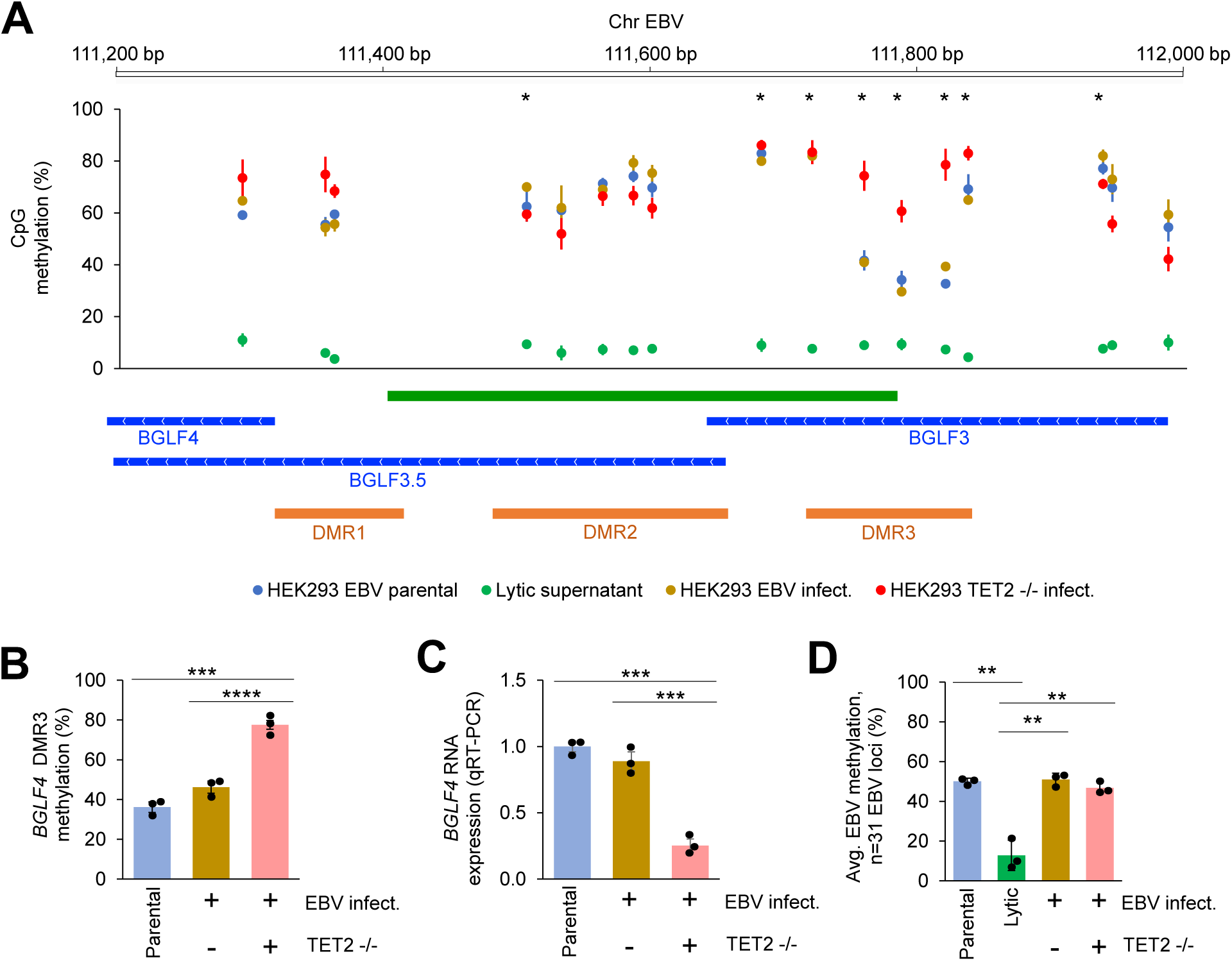
TET2 knockout increases *BGLF4* promoter DNA methylation and lowers *BGLF4* expression. (**A**) Methylation analysis of the *BGLF4* upstream locus using EpiTYPER assay in M81 EBV HEK293 cells (blue: parental, green: lytic induction, brown: EBV infected HEK293 wild type control, red: EBV infected HEK293 TET2−/−). EBV genomic coordinates and statistical significance for individual CpGs are annotated above. Lower panel: CpG islands (green), EBV transcripts (blue) and DMRs (orange) are annotated. (**B,C**) *BGLF4* DNA methylation average from DMR3 CpGs (B, ChrEBV:111757, 111785, 111818) and *BGLF4* RNA expression (C) in parental, wild-type reinfected control or TET2−/− re-infected HEK293 cells. (**D**) EBV DNA methylation average from n=31 EBV CpG sites determined via iPLEX assay in parental, lytic induced, re-infected or TET2−/− re-infected HEK293 cells. Data show mean±standard deviation of biological triplicates. *p<0.05, **p<0.01, ***p<0.001, ****p<0.0001, t-test (Fig 4A: Exact Wilcoxon-Mann-Whitney test).

## Discussion

EBV+ CNSL is a unique disease entity with poor treatment responses and few dedicated treatment options (36). Lymphoma-associated EBV is believed to be strictly latent and silenced most of its genes. Thus, previous research has almost exclusively focused on interventions that re-activate EBV and make it vulnerable to antiviral treatments or immune detection (5,6). Here we provide further evidence that antivirals, like the GARD regimen, can have efficacy in EBV-associated CNSL without EBV re-activation. We show a superior response rate, 2-year survival, and TRM compared to similar CNLS cohorts (11,37,38) that assessed chemotherapy, including methotrexate, and radiation therapy.

EBV lytic activity has so far been considered a switch-like cascade of involving dozens of viral lytic genes expressed (2). Meanwhile, there is evidence that lytic genes have activities outside of full virus activation (39), including *BGLF4* (40). It has, however, remained unclear how a partial EBV activation state – termed abortive lytic cycle (41) – can be achieved. Early findings on the expression of *BGLF4* indicated that *BGLF4* production required viral DNA replication, and thus passive EBV methylation loss (42). *BGLF4* was subsequently shown to be also expressed independently of viral replication (30), and we suggest that DNA methylation loss in this setting may be carried out by TET2. Identifying the determinants of *BGLF4* expression is clinically important, as its expression is mechanistically linked to GCV susceptibility (16). Here, we report experimental evidence that *BGLF4* is undergoing specific epigenetic activation through DNA methylation loss. Rather than being part of a global EBV activation program, we provide evidence that *BGLF4* promoters are switched on in a locus-specific manner, thus challenging the notion of a lytic/latent virus switch and instead proposing a more nuanced, context-dependent regulation of this EBV locus.

With an *in vitro* model of latency establishment, we establish that TET2 is required for large parts of *BGLF4* expression. TET2 has been shown to have crucial functions in EBV genomic activity (34,43). We expand on these findings by establishing the specific requirement of TET2 for *BGLF4* methylation decreases associated with its expression in PTLD and CNSL. Of note, TET2 and IDH1/2 mutations are enriched in EBV+CNSL and other EBV-linked lymphomas (44–46). Based on our *in vitro* models, we propose that such mutations hold significance for EBV’s biological state and may be linked to altered susceptibility to antiviral drugs by affecting *BGLF4* expression. The increase in *BGLF4* luciferase reporter activity in HEK293 cells carrying EBV (Suppl. Fig 4A) points to the contribution of viral factors to *BGLF4* promoter activity.

While the EBV life cycle has been studied in great detail, most of this knowledge has not been applied to clinical care, where the only available diagnostic assay is EBV load detection via PCR. High tumor load in EBV malignancy can mimic the high viral loads normally associated with lytic activation, making these two clinically distinct states indistinguishable via EBV qPCR (8,9). Even in the absence of malignancy, approximately 20% of high-risk individuals show detectable EBV in the CNS (47). We propose that EBV methylation can be leveraged to characterize EBV clinically and derive therapeutic benefit through more informed use of treatments like antivirals. In CNSL patients, EBV is frequently detectable in cerebrospinal fluid (CSF), offering a minimally invasive way to detect EBV from CSF DNA (8,48). The viral DNA methylation state could be equally read out in CSF using DNA methylation assays. We identified key methylation sites associated with *BGLF4* expression, thus setting the stage for biomarker studies utilizing *BGLF4* promoter methylation. In the future, this may offer an opportunity to screen for non-methylated *BGLF4* and initiate targeted antiviral treatment of CNSL.

## Data Availability

All data produced in the present work are contained in the manuscript

## Acknowledgements

We would like to thank the OSU Tissue Archive Services for sample procurement and distribution. We thank Betsy Pray (OSU Dept. Hematology) for support with MaxCyte electroporation procedures. We thank Henri-Jacques Delecluse’s group for providing plasmids and cell lines for EBV M81 virus experiments and Matthew Purcell (OSU Experimental Hematology Laboratory) for assistance with cell sorting. Haley Klimaszewski was supported by the American Society of Hematology (ASH) Medical Student Physician-Scientist Award.

## References

1. Damania B, Kenney SC, Raab-Traub N. Epstein-Barr virus: Biology and clinical disease. Cell 2022;185:3652–70

2. Buschle A, Hammerschmidt W. Epigenetic lifestyle of Epstein-Barr virus. Semin Immunopathol 2020;42:131–42

3. Arvey A, Tempera I, Tsai K, Chen HS, Tikhmyanova N, Klichinsky M, et al. An atlas of the Epstein-Barr virus transcriptome and epigenome reveals host-virus regulatory interactions. Cell Host Microbe 2012;12:233–45

4. Woellmer A, Hammerschmidt W. Epstein-Barr virus and host cell methylation: regulation of latency, replication and virus reactivation. Curr Opin Virol 2013;3:260–5

5. Dalton T, Doubrovina E, Pankov D, Reynolds R, Scholze H, Selvakumar A, et al. Epigenetic reprogramming sensitizes immunologically silent EBV+ lymphomas to virus-directed immunotherapy. Blood 2020;135:1870–81

6. Roychowdhury S, Peng R, Baiocchi RA, Bhatt D, Vourganti S, Grecula J, et al. Experimental treatment of Epstein-Barr virus-associated primary central nervous system lymphoma. Cancer Res 2003;63:965–71

7. Haverkos B, Alpdogan O, Baiocchi R, Brammer JE, Feldman TA, Capra M, et al. Targeted therapy with nanatinostat and valganciclovir in recurrent EBV-positive lymphoid malignancies: a phase 1b/2 study. Blood Adv 2023;7:6339–50

8. Wang Y, Yang J, Wen Y. Lessons from Epstein-Barr virus DNA detection in cerebrospinal fluid as a diagnostic tool for EBV-induced central nervous system dysfunction among HIV-positive patients. Biomed Pharmacother 2022;145:112392

9. Bakker NA, van Imhoff GW, Verschuuren EA, van Son WJ. Presentation and early detection of post-transplant lymphoproliferative disorder after solid organ transplantation. Transpl Int 2007;20:207–18

10. Dharnidharka VR, Webster AC, Martinez OM, Preiksaitis JK, Leblond V, Choquet S. Post-transplant lymphoproliferative disorders. Nat Rev Dis Primers 2016;2:15088

11. Evens AM, Choquet S, Kroll-Desrosiers AR, Jagadeesh D, Smith SM, Morschhauser F, et al. Primary CNS posttransplant lymphoproliferative disease (PTLD): an international report of 84 cases in the modern era. Am J Transplant 2013;13:1512–22

12. Andersen O, Ernberg I, Hedstrom AK. Treatment Options for Epstein-Barr Virus-Related Disorders of the Central Nervous System. Infect Drug Resist 2023;16:4599–620

13. Dugan JP, Haverkos BM, Villagomez L, Martin LK, Lustberg M, Patton J, et al. Complete and Durable Responses in Primary Central Nervous System Posttransplant Lymphoproliferative Disorder with Zidovudine, Ganciclovir, Rituximab, and Dexamethasone. Clin Cancer Res 2018;24:3273–81

14. Grommes C, DeAngelis LM. Primary CNS Lymphoma. J Clin Oncol 2017;35:2410–8

15. Markouli M, Ullah F, Omar N, Apostolopoulou A, Dhillon P, Diamantopoulos P, et al. Recent Advances in Adult Post-Transplant Lymphoproliferative Disorder. Cancers (Basel) 2022;14

16. Meng Q, Hagemeier SR, Fingeroth JD, Gershburg E, Pagano JS, Kenney SC. The Epstein-Barr virus (EBV)-encoded protein kinase, EBV-PK, but not the thymidine kinase (EBV-TK), is required for ganciclovir and acyclovir inhibition of lytic viral production. J Virol 2010;84:4534–42

17. Gustafson EA, Chillemi AC, Sage DR, Fingeroth JD. The Epstein-Barr virus thymidine kinase does not phosphorylate ganciclovir or acyclovir and demonstrates a narrow substrate specificity compared to the herpes simplex virus type 1 thymidine kinase. Antimicrob Agents Chemother 1998;42:2923–31

18. Bayraktar UD, Diaz LA, Ashlock B, Toomey N, Cabral L, Bayraktar S, et al. Zidovudine-based lytic-inducing chemotherapy for Epstein-Barr virus-related lymphomas. Leuk Lymphoma 2014;55:786–94

19. Abrey LE, Batchelor TT, Ferreri AJ, Gospodarowicz M, Pulczynski EJ, Zucca E, et al. Report of an international workshop to standardize baseline evaluation and response criteria for primary CNS lymphoma. J Clin Oncol 2005;23:5034–43

20. Ahmed EH, Lustberg M, Hale C, Sloan S, Mao C, Zhang X, et al. Follicular Helper and Regulatory T Cells Drive the Development of Spontaneous Epstein-Barr Virus Lymphoproliferative Disorder. Cancers (Basel) 2023;15

21. Tsai MH, Lin X, Shumilov A, Bernhardt K, Feederle R, Poirey R, et al. The biological properties of different Epstein-Barr virus strains explain their association with various types of cancers. Oncotarget 2017;8:10238–54

22. Campeau E, Ruhl VE, Rodier F, Smith CL, Rahmberg BL, Fuss JO, et al. A versatile viral system for expression and depletion of proteins in mammalian cells. PLoS One 2009;4:e6529

23. Kim E, Ilic N, Shrestha Y, Zou L, Kamburov A, Zhu C, et al. Systematic Functional Interrogation of Rare Cancer Variants Identifies Oncogenic Alleles. Cancer Discov 2016;6:714–26

24. Giacopelli B, Zhao Q, Ruppert AS, Agyeman A, Weigel C, Wu YZ, et al. Developmental subtypes assessed by DNA methylation-iPLEX forecast the natural history of chronic lymphocytic leukemia. Blood 2019;134:688–98

25. Weigel C, Veldwijk MR, Oakes CC, Seibold P, Slynko A, Liesenfeld DB, et al. Epigenetic regulation of diacylglycerol kinase alpha promotes radiation-induced fibrosis. Nat Commun 2016;7:10893

26. Scotto-Lavino E, Du G, Frohman MA. 5’ end cDNA amplification using classic RACE. Nat Protoc 2006;1:2555–62

27. de Hoon MJ, Imoto S, Nolan J, Miyano S. Open source clustering software. Bioinformatics 2004;20:1453–4

28. Spitzer M, Wildenhain J, Rappsilber J, Tyers M. BoxPlotR: a web tool for generation of box plots. Nat Methods 2014;11:121–2

29. Ehrich M, Nelson MR, Stanssens P, Zabeau M, Liloglou T, Xinarianos G, et al. Quantitative high-throughput analysis of DNA methylation patterns by base-specific cleavage and mass spectrometry. Proc Natl Acad Sci U S A 2005;102:15785–90

30. Djavadian R, Hayes M, Johannsen E. CAGE-seq analysis of Epstein-Barr virus lytic gene transcription: 3 kinetic classes from 2 mechanisms. PLoS Pathog 2018;14:e1007114

31. Wang JT, Chuang YC, Chen KL, Lu CC, Doong SL, Cheng HH, et al. Characterization of Epstein-Barr virus BGLF4 kinase expression control at the transcriptional and translational levels. J Gen Virol 2010;91:2186–96

32. Verma N, Pan H, Dore LC, Shukla A, Li QV, Pelham-Webb B, et al. TET proteins safeguard bivalent promoters from de novo methylation in human embryonic stem cells. Nat Genet 2018;50:83–95

33. Wille CK, Li Y, Rui L, Johannsen EC, Kenney SC. Restricted TET2 Expression in Germinal Center Type B Cells Promotes Stringent Epstein-Barr Virus Latency. J Virol 2017;91

34. Lu F, Wiedmer A, Martin KA, Wickramasinghe P, Kossenkov AV, Lieberman PM. Coordinate Regulation of TET2 and EBNA2 Controls the DNA Methylation State of Latent Epstein-Barr Virus. J Virol 2017;91

35. Cairns RA, Mak TW. Oncogenic isocitrate dehydrogenase mutations: mechanisms, models, and clinical opportunities. Cancer Discov 2013;3:730–41

36. Gandhi MK, Hoang T, Law SC, Brosda S, O’Rourke K, Tobin JWD, et al. EBV-associated primary CNS lymphoma occurring after immunosuppression is a distinct immunobiological entity. Blood 2021;137:1468–77

37. Snanoudj R, Durrbach A, Leblond V, Caillard S, Hurault De Ligny B, Noel C, et al. Primary brain lymphomas after kidney transplantation: presentation and outcome. Transplantation 2003;76:930–7

38. Cavaliere R, Petroni G, Lopes MB, Schiff D, International Primary Central Nervous System Lymphoma Collaborative G. Primary central nervous system post-transplantation lymphoproliferative disorder: an International Primary Central Nervous System Lymphoma Collaborative Group Report. Cancer 2010;116:863–70

39. Munz C. The Role of Lytic Infection for Lymphomagenesis of Human gamma-Herpesviruses. Front Cell Infect Microbiol 2021;11:605258

40. Li R, Liao G, Nirujogi RS, Pinto SM, Shaw PG, Huang TC, et al. Phosphoproteomic Profiling Reveals Epstein-Barr Virus Protein Kinase Integration of DNA Damage Response and Mitotic Signaling. PLoS Pathog 2015;11:e1005346

41. Munz C. Tumor Microenvironment Conditioning by Abortive Lytic Replication of Oncogenic gamma-Herpesviruses. Adv Exp Med Biol 2020;1225:127–35

42. Gershburg E, Marschall M, Hong K, Pagano JS. Expression and localization of the Epstein-Barr virus-encoded protein kinase. J Virol 2004;78:12140–6

43. Wille CK, Nawandar DM, Henning AN, Ma S, Oetting KM, Lee D, et al. 5-hydroxymethylation of the EBV genome regulates the latent to lytic switch. Proc Natl Acad Sci U S A 2015;112:E7257–65

44. Cho J, Kim E, Yoon SE, Kim SJ, Kim WS. TET2 and LILRB1 mutations are frequent in Epstein-Barr virus-positive diffuse large B-cell lymphoma especially in elderly patients. Cancer 2023;129:1502–12

45. Radke J, Ishaque N, Koll R, Gu Z, Schumann E, Sieverling L, et al. The genomic and transcriptional landscape of primary central nervous system lymphoma. Nat Commun 2022;13:2558

46. Hayashi T, Tateishi K, Matsuyama S, Iwashita H, Miyake Y, Oshima A, et al. Intraoperative Integrated Diagnostic System for Malignant Central Nervous System Tumors. Clin Cancer Res 2024;30:116–26

47. Musukuma-Chifulo K, Siddiqi OK, Chilyabanyama ON, Bates M, Chisenga CC, Simuyandi M, et al. Epstein-Barr Virus Detection in the Central Nervous System of HIV-Infected Patients. Pathogens 2022;11

48. Bossolasco S, Falk KI, Ponzoni M, Ceserani N, Crippa F, Lazzarin A, et al. Ganciclovir is associated with low or undetectable Epstein-Barr virus DNA load in cerebrospinal fluid of patients with HIV-related primary central nervous system lymphoma. Clin Infect Dis 2006;42:e21–5

