## Supplemental Tables and Figures for "Epigenetic Activation of the EBV Protein Kinase Determines Antiviral Drug Response in Central Nervous System Lymphoma"

**Suppl. Table 1. Comparison of CNSL cases to peripheral PTLD controls.** For age Wilcoxon signed-rank test was used because distribution was non-normal. For sex and age, Fisher exact test was used.

| | | CNSL (n=12) | Peripheral PTLD (n=24) | p-value |
|---|---|---|---|---|
| | | Mean±SD | Mean±SD | |
| **Age** | | 54.6±19) | 54.1±15 | 0.907 |
| | | n= (%) | n= (%) | |
| **Sex** | | | | |
| | Female | 0 (0) | 7 (29) | 0.070 |
| | Male | 12 (100) | 17 (71) | |
| **Race** | | | | |
| | White | 10 (83) | 23 (96) | 0.253 |
| | Indian | 1 (8) | 0 (0) | |
| | Black | 1 (8) | 0 (0) | |
| | Multiple | 0 (0) | 1 (4) | |

### Suppl. Table 2. Oligonucleotide primers and guideRNA. *phosphorothioate-modified oligo

| Primer ID | Forward | Reverse | Comment/details |
|---|---|---|---|
| **qRT-PCR primers (RNA expression analysis)** | | | |
| BXLF1 | GCATAGTAGTCTTTCCCACACC | TCC AAT TGT GAC CCA CCT AAC | Hydrolysis probe: /56FAM/AG GGA ACA G/ZEN/G CAG GTT TGA TTA CTG G/3IABkFQ/ |
| BGLF4 | CCCACATGGTGTCTGTGAAAT | GGCCTCAAAGATGCCGTTTA | Hydrolysis probe: /56-FAM/TA ATG TCC G/ZEN/A ATG GAA GAG GCC GC/3IABkFQ/ |
| LMP1 | TGTCCGTGCAAATTCCAGAG | ACTTGGAGCCCTTTGTATACTC | Hydrolysis probe: /56-FAM/ATCACCCTCCTGCTCATCGCTC/3IABkFQ/ |
| BZLF1 | GTTGCTTAAACTTGGCCCG | ACCACAACAGCCAGAATCG | Hydrolysis probe: /56-FAM/TCTAGTTCAGAATCGCATTCCTCCAGC/3IABkFQ/ |
| ACTB | (see IDT ref. Hs.PT.39a.22214847) | (see IDT ref. Hs.PT.39a.22214847) | (IDT ref. Hs.PT.39a.22214847) |
| **qPCR primers (EBV load determination)** | | | |
| EBNA1 | TCATCATCATCCGGGTCTCC | CCTACAGGGTGGAAAAATGGC | |
| ACTB | CAGGCAGCTCGTAGCTCTTC | TCGTGCGTGACATTAAGGAG | |
| ***TET2* guideRNA and homology templates.** | | | |
| TET2 gRNA | CAGGACTCACACGACTATTC | | |
| TET2 TrueTag template* | TATTATGGAATACCCTGTATGAAGGGA AGCCAGAATGGAAGTGGCTCAGGTTC TGGA | CTACTTTCTTGTGTAAAGTCAGGACTCA CACGACTCTTGGCCGATCGCATACAGA G | |
| **Luciferase reporter assay primers. Entries correspond to reporters shown in Fig. 3A, top to bottom, with all forward primers carrying 5'-AAGCTT(HindIII and all reverse primers 5'-CCTGCAGG(SbfI)** | | | |
| BGLF4_LUCIA_F1R4 | GTTCCTCAAATGGCTCGAGG | CCCCCGTTTACCAGAAGAAT | ChrEBV:111327-112388 |
| BGLF4_LUCIA_F1R3 | GTTCCTCAAATGGCTCGAGG | GCGGTTTATACACCATGTGC | ChrEBV:111327-112063 |
| BGLF4_LUCIA_F1R2 | GTTCCTCAAATGGCTCGAGG | GGAAGCCTCCACTGGAACTA | ChrEBV:111327-111740 |
| BGLF4_LUCIA_F4R4 | TGGTGTATAAACCGCAGTGG | CCCCCGTTTACCAGAAGAAT | ChrEBV:112049-112388 |
| BGLF4_LUCIA_F3R4 | GGAGGCTTCCCGGTAATTT | CCCCCGTTTACCAGAAGAAT | ChrEBV:111731-112388 |
| BGLF4_LUCIA_F1R1 | GTTCCTCAAATGGCTCGAGG | TTTGAAACACAGTTTACATCTGCTC | ChrEBV:111327-111555 |
| BGLF4_LUCIA_F2R4 | CCGAGCAGATGTAAACTGTGTT | CCCCCGTTTACCAGAAGAAT | ChrEBV:111531-112388 |
| BGLF4_LUCIA_F2R3 | CCGAGCAGATGTAAACTGTGTT | GCGGTTTATACACCATGTGC | ChrEBV:111531-112063 |
| BGLF4_LUCIA_F2R2 | CCGAGCAGATGTAAACTGTGTT | GGAAGCCTCCACTGGAACTA | ChrEBV:111531-111740 |
| BGLF4_LUCIA_F3R3 | GGAGGCTTCCCGGTAATTT | GCGGTTTATACACCATGTGC | ChrEBV:111731-112063 |
| **5'RACE primers** | | | |
| 5'RACE_Qt | CCAGTGAGCAGAGTGACGAGGACTCGAGCTCAAGCTTTTTTTTTTTTTTTTT | | |
| 5'RACE_Qo | CCAGTGAGCAGAGTGACG | | |
| 5'RACE_Qi | GAGGACTCGAGCTCAAGC | | |
| BGLF4_GSP_RT1 | CTCAACTCCGCAGCCATATT | | |
| BGLF4_GSP_RT2 | GTGGCGCAGGGACTCTAAG | | |
| BGLF4_GSP1 | CACATCCATGTTCCTCAAATG | | |
| BGLF4_GSP2 | CCCGAGCAGATGTAAACTGT | | |
| BGLF4_GSP3 | CTGTCGCAGCTCGTCTCT | | |
| BGLF4_GSP4 | GTTTCAAACAGAGCGTGGTC | | |
| **EpiTYPER methylation analysis primers. All forward primers carry 5'tag AGGAAGAGAG, all reverse primers carry 5'tag CAGTAATACGACTCACTATAGGGAGAAGGCT** | | | |
| BGLF4_1 | AGGGGGTTTTGGGGAGATATTTA | CCAAAAATCAACTACAAACAACTAAAA | ChrEBV:111246-111429 |
| BGLF4_2 | GGTGATTATTTTTTGATAGGTAGAAA | TCTAATAACCCAACAACTAAAAACCTC | ChrEBV:111448-111640 |
| BGLF4_3 | GTGTGTGTTTTTTGTGTTTGTTAT | CTCATCTTCATAAATCACCATATCC | ChrEBV:111656-111876 |
| BGLF4_4 | GTGATGTAAAAGGGTTTTTGAGGTA | CATTCCTAACACACCTCACCAAATA | ChrEBV:111879-112032 |
| BZLF1 | GGTTTGATTGGTTTTTTTATTAGGG | ACCCCTACCTACCTCTTTAACTCC | ChrEBV:91358-91520 |
| BXLF1 | GTTGTGTGATTGTGTTAATTTTTTT | TTTTAACCCAAAACTAAAAACTCTACCT | ChrEBV:132930-133073 |
| LMP1 | GGTGGTTAAGTGTAATAGGAAATGG | ATTACCCCACAACCTTACCTCACCTA | ChrEBV:169258-169471 |
| LINE1 | TTTATATTTTGGTATGATTTTGTAG | TTTATCACCACCAAACCTACCCT | |
| Alu-e | GTTTGTAATTTTAGTATTTTGGGAGG | CCTACCTCAACCTCCCRAATAA | |
| LTR1 | TAAGGTAGAGGGTTTAATTGAGTTGG | AAATAAAAAAAAATTCCCTTATCCCC | |
| **Methylation iPLEX primers. All forward and reverse primers carry 5'tag ACGTTGGATG. Lowercase bases: oligo mass tags, non-EBV sequence** | | | |
| BHRF1 | GAGTTGTGTTTTAGGTTTGTG | CCACAAACCCCAATACGTAA | multiplex PCR 1, ext. oligo: cTAGGTTTGTGTTTTGTTTTG |
| BMRF1 | TTATCGAGGTTCGTGGAGAG | CACCCAAAATACGATCGACC | multiplex PCR 1, ext. oligo: GGTTATTTTGGTGGATGTG |
| BORF1 | TATAGCGGACGTTATGAAGG | AAACAACAACCCCGCGATTC | multiplex PCR 1, ext. oligo: ccTTATGAAGGTTTAGGGGTT |
| BVRF1 | CGGGAATATTAATTAGTGGG | CCCCTCAACTACTTAAAAAAC | multiplex PCR 1, ext. oligo: ttGGATTTTGGTATAGGATTATGT |
| EBER1 | GGGTTTCGGAGTTTTTAGG | CACCCAACACTACACGTAAC | multiplex PCR 1, ext. oligo: aGTTAGTTTGTAAGGTGGATGG |
| LMP1 | TAGGTGAGGTAAGGTTGTGG | CGACCCTTAACCCTCTTAAC | multiplex PCR 1, ext. oligo: ggGAGGTAAGGTTGTGGGGTAAT |
| LMP2B | TAGAGAGCGATGAGTAGGAG | CCCTTTCCCTTACTCTTCC | multiplex PCR 1, ext. oligo: aGGGGAAAGAGGAGAAAGTG |
| Rp | GTTTAAGTATGAGTGGGTAG | CCAACCAAATATTCAAAAACC | multiplex PCR 1, ext. oligo: GAGTGGGTAGTAGAGAGGTT |
| BARF0 | GCGTATTTGCGGCGTTATAG | ACACCTTCATCACGAAACCC | multiplex PCR 2, ext. oligo: tgGTTGGGTGGGGGAAAGAGTT |
| BFRF1 | AGTTGAGATCGAAGACGAGG | AATTCCTTCTTCCACGACGC | multiplex PCR 2, ext. oligo: TTTTTTGGAGGGATAAAGATT |
| BORF1 | GGGGAGTTTTAGTTGGTTT | AACGTTCCAATAACCTCGAC | multiplex PCR 2, ext. oligo: ccccTTTTAGTTGGTTTGGGTGG |
| OriLyt | TCGTCGTAAGGACGTCGGGT | AAAACGAACCGACCCGACTC | multiplex PCR 2, ext. oligo: TGGGAGGTGTGTATTTTT |
| Qp | GGGTGATTATTGAGGGAGTG | TCGCAAAACGTAACCTCACC | multiplex PCR 2, ext. oligo: GTTTTATAGTAATGTTGTTTGGT |
| Wp | CGTATAATGGCGGATTTAGG | CTTATTCCTCTTTTCCCCTC | multiplex PCR 2, ext. oligo: GGGTTTATGGTTGGTTG |
| BSRF1 | TGGTTGTAAGYGGTATYGAG | TCTCCATCTCCATCAAAACC | multiplex PCR 3, ext. oligo: cgtgTTGTTGGATAAATAGGTTGG |
| EBNA1 | TYGYGTAGGTTTTTTTTAGG | TCACCATCTAAACCACCTTC | multiplex PCR 3, ext. oligo: TTTTTTTATTTTGTAGGGGAAGT |
| LF3 | TGTTTTTTTGGGTTTTAGG | CCCRGCRGAAARGAAACAA | multiplex PCR 3, ext. oligo: TTAGGATTTAGTTTTGGAGTT |
| LMP2A/B | TYGGYGGAGAGAAYGTTAAG | TACCAACCTAATTCCCTCCC | multiplex PCR 3, ext. oligo: gggaGAGAATAGGGGTATAAAGTGT |
| BLLF1 | GTTGTTGTGGTGGTATTGGTAG | RGAACATTTAACATTACTATCTC | multiplex PCR 4, ext. oligo: ATGAGTGTTTTGGGGGT |
| EBER1 | YGTTGTTTTAGAGGTTTTGTTAG | ACCRGAAACTTATACCRGAAAC | multiplex PCR 4, ext. oligo: tGTTTTGTTAGGGAGGAGA |
| BRLF1 | AGGAAGAAAGGTGGGTTTTGAG | AARGRGARGAAACCTAAACAAC | multiplex PCR 4, ext. oligo: GGGGAATATGGGTTTTTTAT |
| EBNA1 | GGGTGATGAAAATAGAGGTTGG | AAATAACRGAACAAAACAACCRG | multiplex PCR 4, ext. oligo: GAAAATAGAGGTTGGATGTAG |
| LMP2A | GAGGGAATTAGGTTGGTAATGG | TAAACAAAARGTCTTCRGAATCC | multiplex PCR 4, ext. oligo: ggAGGTTGGTAATGGTTGAGG |
| BZLF1 | TGTTTGTTAGTTGTTGTTTTTGG | CCTCAACCTAAAAACAATTCTAC | multiplex PCR 4, ext. oligo: AGTTGTTGTTTTTGGTTAGTTT |
| Cp | GTGGGAAAAAAATTTATGGT | TTATTCCTTCTCCcrCCAAC | multiplex PCR 4, ext. oligo: GGGAAAAAAATTTATGGTTTAGTG |
| BALF2 | TAGYGTTTGGTAGTGATTTTTTAG | TCTCCCTACCCATCTATATATAC | multiplex PCR 5, ext. oligo: GGGGGTTTTGGTTAAGTT |
| LMP1 | AAAGTTGTTGGAGTTATTGGGTG | AACCCCTTACTTCCACAAAATAC | multiplex PCR 5, ext. oligo: tGGGATGTATGTGGTTTTT |
| BMRF1 | AGTTAAGGTTGATYGATTAAGGG | CRATAAAAACAATCTCTACCTCC | multiplex PCR 5, ext. oligo: tTTTGGATTTAGGAGTTGTTT |
| BFLF1/2 | TATTTGTGTTGAAGATGGGGTAG | AAACTAAACTTTCCCAACCCAAC | multiplex PCR 5, ext. oligo: AGATGGTGTTATAAAAGTGTT |
| BHRF1 | GAAGGTTTGGATGGTTGGATTTA | CCAACTAAACCTTCTAAATCCAAA | multiplex PCR 5, ext. oligo: ccTGGTTGGATTTATTAATAGGG |
| BNLF2a | GAGTATTATTAGGAGTAGTTTTAG | CTTCTACRAAAACCAAACCTAAC | multiplex PCR 5, ext. oligo: gcGGAGTAGTTTTAGTTTGTTGA |

**Suppl. Table 3. *BGLF4* transcription start sites (TSS) determined via 5'RACE.** Table shows cell line/sample name, EBV genome coordinate (RefSeq ID NC_007605.1) of detected TSS, and PCR primers used in 5'RACE reverse transcription (RT) and two nested PCRs. [1]forward primer: 5'RACE_Qo, [2]: forward primer: 5'RACE_Qi

| Sample | TSS (ChrEBV) | GSP-RT anchor primer | primer (PCR1)[1] | primer (PCR2)[2] |
|---|---|---|---|---|
| Raji | N/A | GSP_RT1 | GSP_1 | GSP_3 |
| Rael | 111527 | GSP_RT1 | GSP_1 | GSP_3 |
| LCL EBV B95-8 | 111581 | GSP_RT1 | GSP_1 | GSP_3 |
| LCL EBV B95-8 | 111545 | GSP_RT1 | GSP_1 | GSP_3 |
| LCL EBV B95-8 | 111793 | GSP_RT2 | GSP2 | GSP4 |
| HEK293 EBV | 111793 | GSP_RT2 | GSP2 | GSP4 |
| CNSL #1 | 111527 | GSP_RT1 | GSP_1 | GSP_3 |
| CNSL #1 | 111793 | GSP_RT2 | GSP2 | GSP4 |
| CNSL #2 | 111793 | GSP_RT2 | GSP2 | GSP4 |
| CNSL #3 | 111527 | GSP_RT1 | GSP_1 | GSP_3 |
| CNSL #4 | 111790 | GSP_RT2 | GSP2 | GSP4 |
| CNSL #4 | 111581 | GSP_RT1 | GSP_1 | GSP_3 |
| CNSL #4 | 111527 | GSP_RT1 | GSP_1 | GSP_3 |
| CNSL #5 | 111568 | GSP_RT1 | GSP_1 | GSP_3 |
| CNSL #5 | 111581 | GSP_RT1 | GSP_1 | GSP_3 |

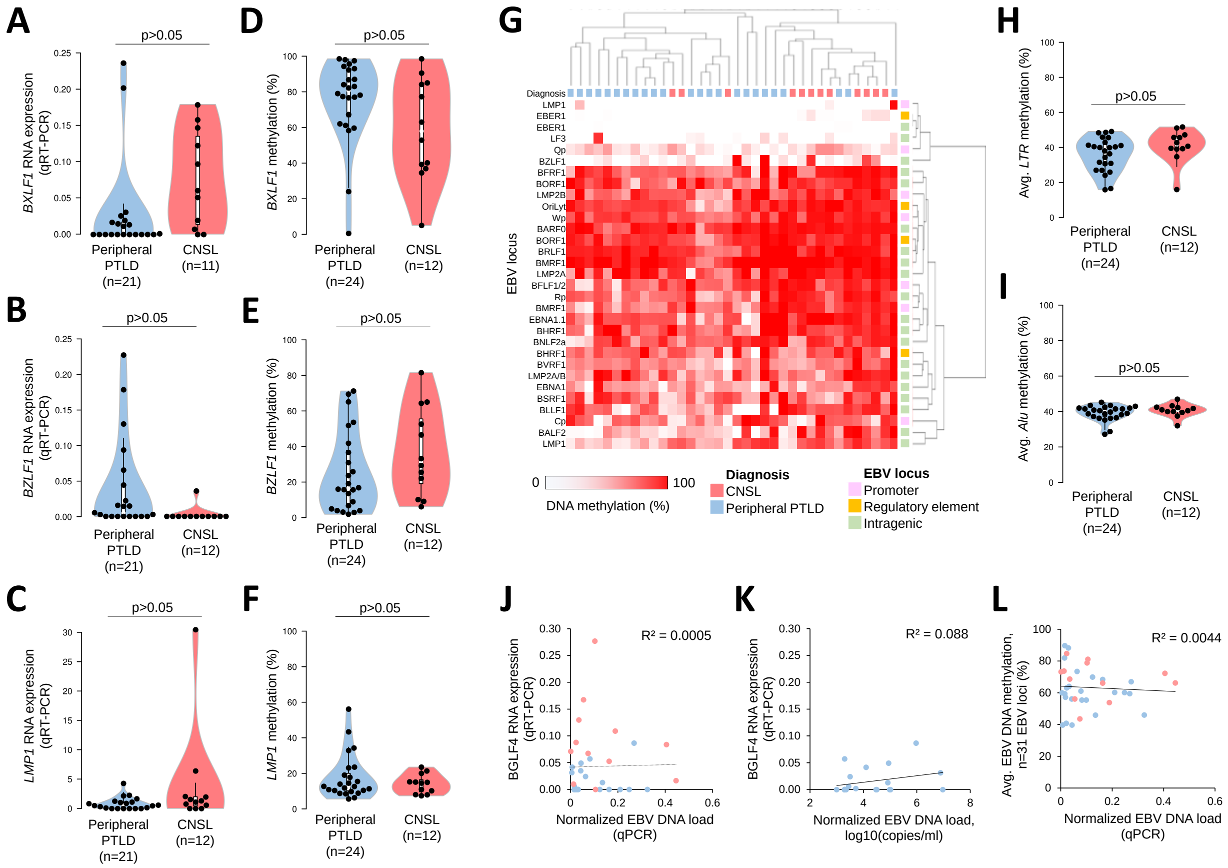


**Suppl. Figure 2. EBV DNA methylation and gene expression in CNSL.** (**A-C**) RNA expression (qRT-PCR) and (**D-F**) DNA methylation determined via EpiTYPER in peripheral PTLD and CNSL tissue biopsies for EBV *BXLF1* (A,D), *BZLF1* (B,E) and *LMP1* (C,F). (**G**) Heatmap depicting EBV DNA methylation from n=31 methylation sites determined with methylation iPLEX assay. Samples were arranged via unsupervised clustering and annotated based on sample cohort and EBV locus. DNA methylation is depicted from 0% methylated (white) to 100% methylated (red). (**H,I**) Average DNA methylation of human repetitive elements *LTR* (H) and *Alu* (I) determined via EpiTYPER in CNSL (n=12) and peripheral PTLD (n=24) cohorts. (**J,K**) correlation of *BGLF4* RNA expression and EBV DNA load in determined via qPCR tissue samples (J) or peripheral blood EBV DNA load from clinical chart review (K). (**L**) correlation of average EBV DNA methylation determined via iPLEX assay (n=31 EBV methylation sites) and EBV load in tissue biopsies determined via qCPR. Sample cohorts are annotated in red (CNSL) and blue (peripheral PTLD). $R^2$: correlation coefficient. *$p<0.05$, t-test (D-F: Exact Wilcoxon-Mann-Whitney test)

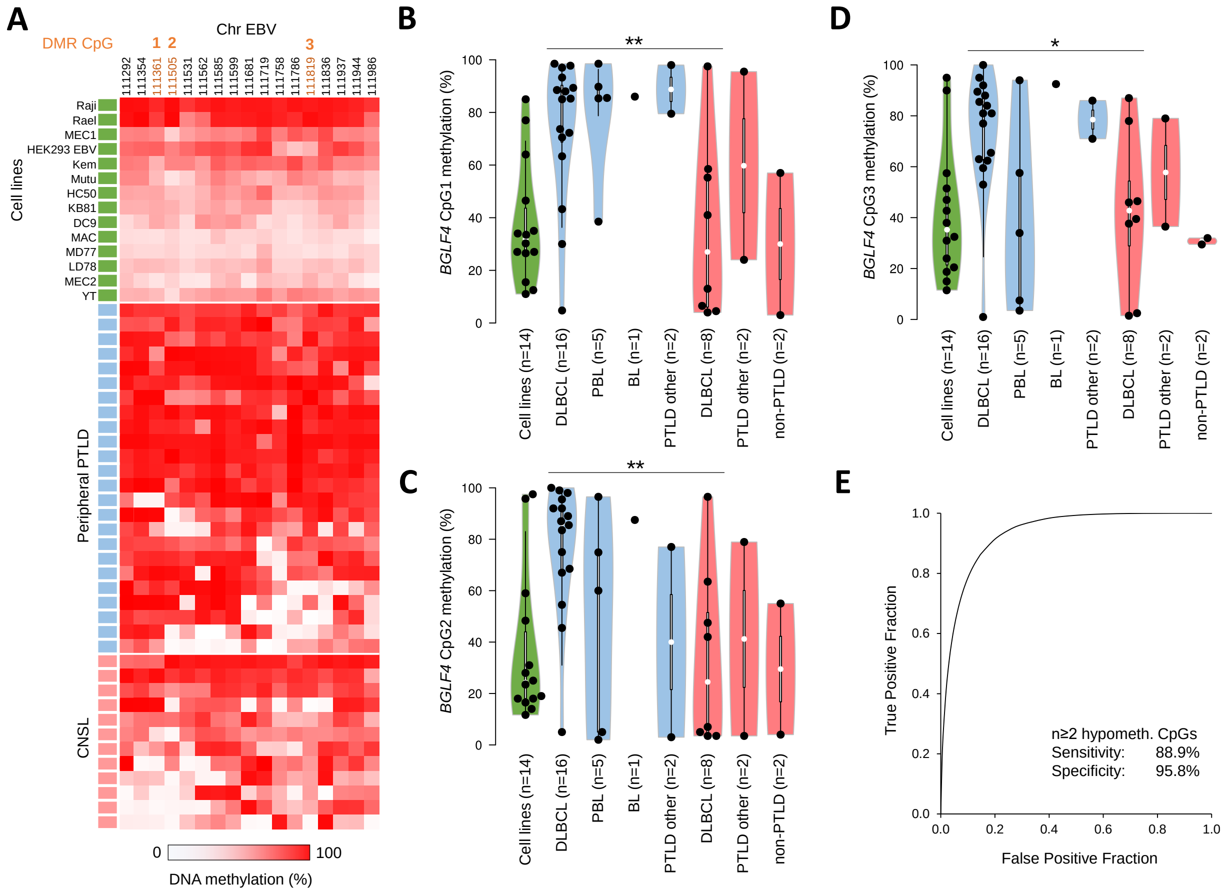


**Suppl. Figure 3. Site-specific methylation loss at EBV *BGLF4*.** (**A**) Heatmap depicting EBV DNA methylation at the *BGLF4* upstream region determined via EpiTYPER assay. Samples were annotated based on sample cohort (green: cell lines, blue: peripheral PTLD, red: CNSL). EBV genome coordinates (EBV genome Refseq ID: NC_007605.1) are shown above, with DMR CpG 1, 2 and 3 annotated in orange. DNA methylation is depicted from 0% methylated (white) to 100% methylated (red). (**B-D**) DNA methylation for DMR CpGs 1 (B), 2 (C) and 3 (D) in cell lines (green), peripheral PTLDs (blue) and CNSL (red). Cohorts were separated based on pathology reports. DLBCL: Diffuse Large B-cell lymphoma, PBL: plasmablastic lymphoma, BL: Burkitt lymphoma. (**E**) ROC curve and sensitivity/specificity for n≥2 hypomethylated (<50% methylation) DMR CpGs as a marker of high *BGLF4* RNA expression (qRT-PCR, $4^{th}$ quartile of combined CNSL/peripheral PTLD cohort). *p<0.05, **p<0.01, t-test.

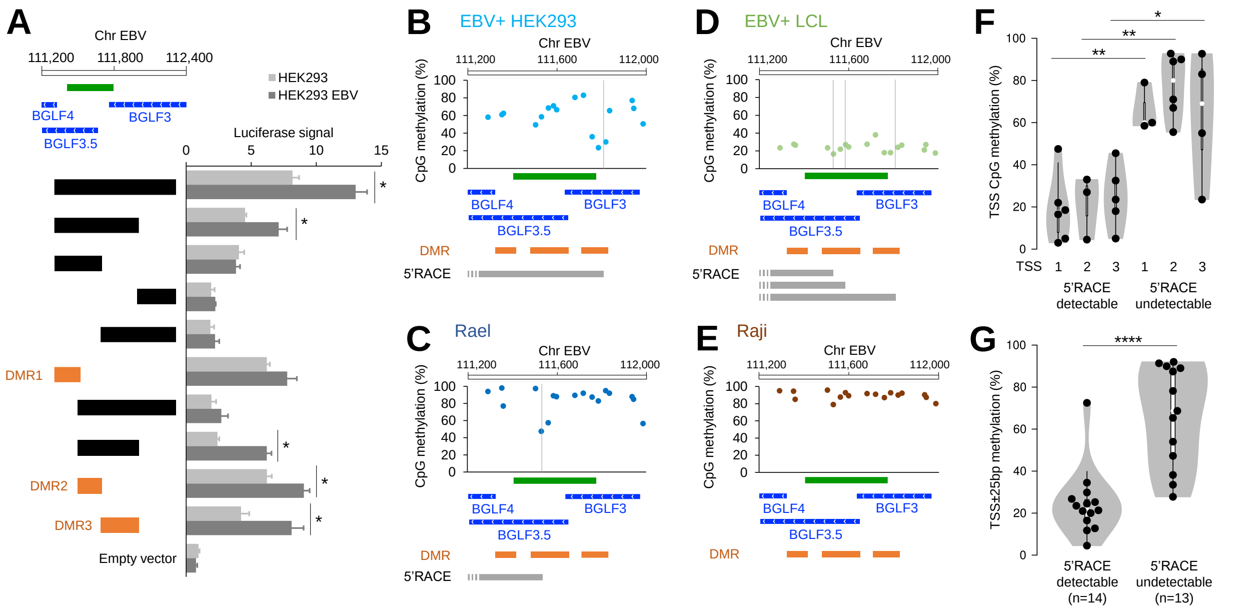


**Suppl. Figure 4. BGLF4 DMRs possess gene promoter activity.** (**A**) Luciferase reporter assay of *BGLF4* upstream DNA elements in HEK293 or HEK293 M81 EBV cells. DNA sequences representing DMRs are marked in orange. EBV genomic coordinates, CpG islands (green) and EBV transcripts (blue) are indicated above. Bar graphs show mean±standard deviation of quadruplicate readouts. (**B-E**) Results from 5'RACE in n=4 cell line samples with detectable *BGLF4* RNA expression. Plots show DNA methylation across the *BGLF4* upstream region in each sample with detected TSS marked by vertical lines. Lower panels: CpG islands (green), EBV transcripts (blue), DMRs (orange) and Sequenced transcripts from 5'RACE (grey). (**F,G**) Violin plots depicting DNA methylation at single CpG sites closest to three TSS from 5'RACE (G) or average CpG methylation of all CpGs within 25 bp around TSS (G) with either detectable (total n=14) or undetectable (total n=13) transcript. TSS1: ChrEBV:111527, TSS2: ChrEBV:111581, TSS3: ChrEBV: 111793). Data are aggregate readouts from n=5 CNSL samples and n=4 EBV+ cell lines. *p<0.05, **p<0.01, ****p<0.0001, t-test (F,G: Exact Wilcoxon-Mann-Whitney test).

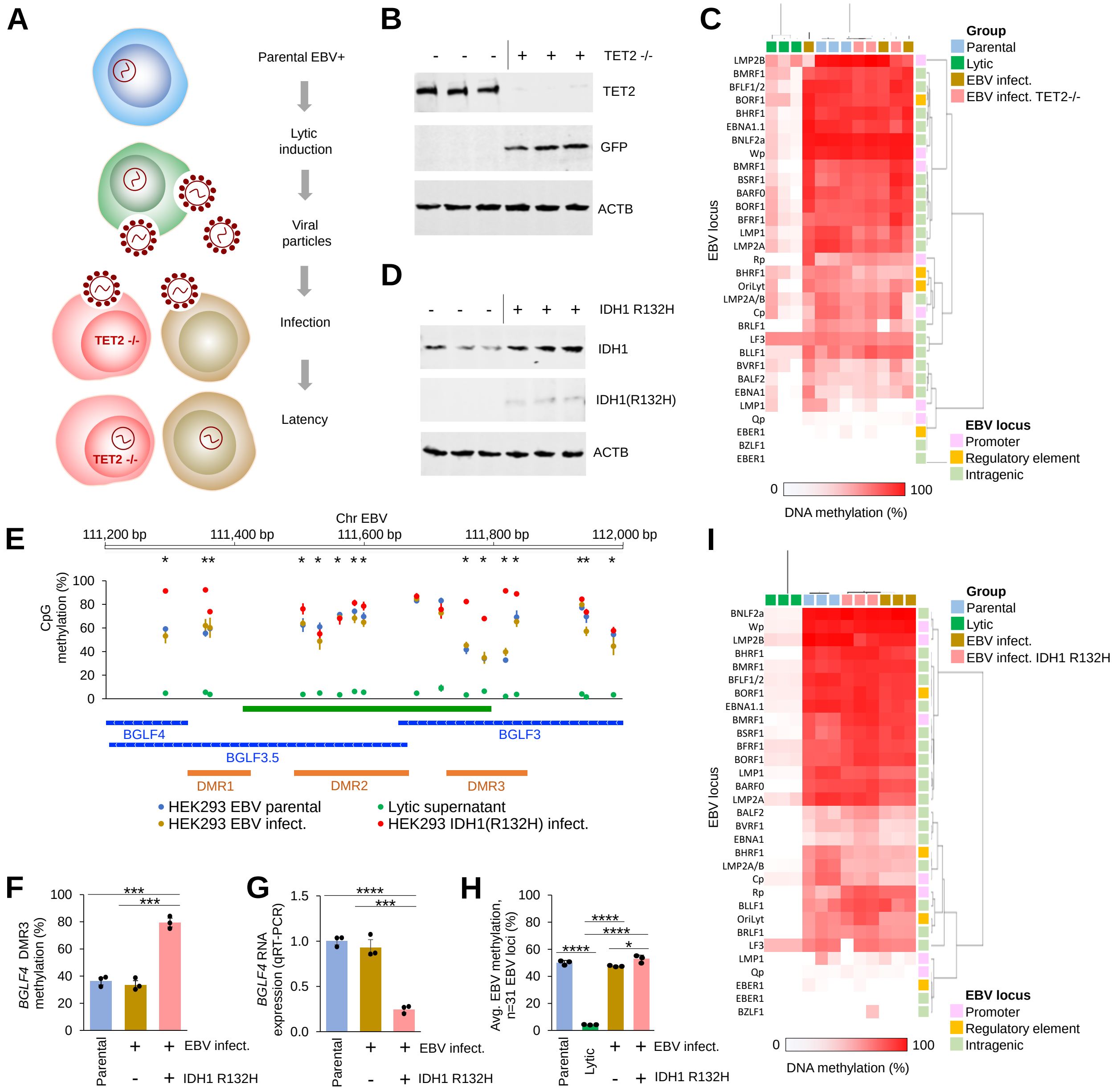

**Suppl. Figure 5. Loss of TET function increases *BGLF4* promoter DNA methylation and lowers *BGLF4* expression.** (**A**) Schematic of EBV latency establishment experiments. Parental M81 EBV HEK293 cells (blue) undergo lytic induction via lytic gene introduction (green). The collected viral particles are then used to infect HEK293 cells (either wild type, brown or TET2-/-, red) and latency establishment occurs. (**B**) Western Immunoblot showing knockout of TET2 in CRISPR-Cas9 modified HEK293 cells and simultaneous knock-in of green fluorescent protein (GFP). Triplicate experiments are shown. (**C**) Heatmap depicting EBV DNA methylation from n=31 methylation sites determined via methylation iPLEX assay in parental M81 EBV HEK293 (blue), after lytic induction (green), EBV-infected HEK293 wild type control (brown) or TET2-/- (red) cells. Samples were arranged via unsupervised clustering and annotated based on sample cohort and EBV locus. (**D**) Western Immunoblot showing stable lentiviral expression of IDH1 or mutated IDH1 R132H in HEK293 cells. Triplicate experiments are shown. (**E**) Methylation analysis of the *BGLF4* upstream locus using EpiTYPER assay in M81 EBV HEK293 cells (blue: parental, green: lytic induction, brown: EBV infected HEK293 wild type control, red: EBV infected HEK293 with lentiviral IDH1 R132H expression). EBV genomic coordinates and statistical significance for individual CpGs are annotated above. Lower panel: CpG islands (green), EBV transcripts (blue) and DMRs (orange). (**F,G**) *BGLF4* DNA methylation average from DMR3 CpGs (F, ChrEBV:111757, 111785, 111818) and *BGLF4* RNA expression (G) in parental (blue), wildtype EBV infected control (brown) or IDH1 R132H-expressing EBV infected (red) HEK293 cells. (**H**) EBV DNA methylation average from n=31 EBV CpG sites determined via iPLEX assay in parental, lytic induced, re-infected or TET2-/- re-infected HEK293 cells. (**I**) Heatmap depicting EBV DNA methylation from n=31 methylation sites determined via methylation iPLEX assay in parental M81 EBV HEK293 (blue), lytic induction (green), EBV infected HEK293 wild type (brown) or EBV infected IDH1 R132H-expressing (red) cells. Samples were arranged via unsupervised clustering and annotated based on sample cohort and EBV locus. DNA methylation in heatmaps is depicted from 0% (white) to 100% methylated (red). All data show mean±standard deviation of biological triplicates. *p<0.05, ***p<0.001, ****p<0.0001, t-test (Suppl. Fig. 4E: Exact Wilcoxon-Mann-Whitney test.).
